# Integrated metabolomics and genetic analyses reveal loss of protective docosahexaenoic acid as a key driver linking ultra-processed food to Crohn’s disease risk

**DOI:** 10.64898/2026.02.20.26346727

**Authors:** Sidan Wang, Lintao Dan, Xixian Ruan, Judith Wellens, Yuhao Sun, Jialu Yao, Li Tian, Rahul Kalla, Evropi Theodoratou, Shuai Yuan, Susanna C Larsson, Jonas F Ludvigsson, Laurent Peyrin-Biroulet, Jack Satsangi, Fernando Magro, Xue Li, Xiaoyan Wang, Jie Chen

## Abstract

**Objectives:** To characterize ultra-processed food (UPF) circulating metabolic signatures associated with Crohn’s disease (CD) and to localize key metabolic mediators linking UPF intake to CD risk.

**Design:** Prospective cohort study.

**Setting:** Two large multi-center cohorts (UK Biobank [UKB] and Whitehall II [WHII] study) across the UK and an Eastern multi-center cohort ONE-IBD Study from China.

**Participants:** UK Biobank discovery cohort (n=10,229) for signature derivation, internal validation cohort (n=91,306), external validation cohort Whitehall-II (n=7,893), and three additional cohorts (two Western and ONE-IBD) for validation of key metabolic drivers.

**Main outcome measures:** Primary outcomes were UPF-related circulating metabolic signatures and their associations with CD risk; secondary outcomes included evidence supporting causal roles of candidate metabolites and genetic pathways assessed by Mendelian randomization, colocalization, and gene-environment analysis.

**Results:** A UPF metabolic signature of 73 metabolites was constructed and validated across cohorts (Spearman ρ: 0.20-0.25). More pronounced UPF metabolic signature was associated with increased CD risk (HR_per SD_=2.65, 95% CI 1.57-4.48). WGCNA revealed a cluster enriched in fatty acids. Within this cluster, docosahexaenoic acid (DHA) emerged as the strongest, which mediated 17.1% of the UPF-CD association. External validation in ONE-IBD supported DHA as the strongest associated metabolite with UPF and CD. Mendelian randomization supported a causal protective effect of DHA on CD (OR=0.72, 95% CI 0.61– 0.83; P<0.001), with colocalization implicating rs174546 in the *FADS1* gene.

**Conclusion:** The adverse effects of UPF on CD risk may be driven by a relative deficiency of protective metabolites such as DHA, apart from additive harm to metabolic depletion. This reframes UPF-related risk and highlighting potential targets for precision nutrition in CD prevention.

## Introduction

Inflammatory bowel disease (IBD), including two major subtypes, Crohn’s disease (CD) and ulcerative colitis (UC), is a lifelong gastrointestinal disease characterized by chronic inflammation with relapsing and remitting phases. Over the past decades, the incidence of IBD, particularly CD, has been rising since the era of global industrialization. This trend is potentially caused by multiple environmental factors, among which diet may be a key player, according to recent data^1^. The transition to the Westernized diet featuring overconsumption of ultra-processed food (UPF) has attracted scrutiny^2^. UPFs are defined as items that undergo a series of processing steps containing energy-dense products high in fat and sugar, and low in dietary fiber, vitamins, and minerals^3^. A detrimental effect of UPF on CD development has been recently noted in one population-based study and synthesized evidence^3^ ^4^. However, human-derived evidence remains scarce on the underlying mechanisms and potential modulators of this association.

Higher UPF intake is reported to induce metabolic perturbations that include alterations in lipid profiles and fatty-acid composition^2^. While UPF-related metabolic signatures have been associated with chronic disease^5^, there is insufficient evidence to link such signatures to incident CD. In the present study, we first developed a metabolic signature associated with UPF intake based on nuclear magnetic resonance (NMR) metabolomic data and investigated its association with incident CD. Subsequently, we applied a systematic, stepwise analytical method to explore key metabolite clusters and specific metabolites in the UPF metabolic signature. We further investigated potentially causal evidence for metabolites prioritized through this framework using Mendelian randomization (MR). Colocalization analysis and gene-environment analysis were used to explore potential mechanisms of genetic variants and their relevance as targets for modulating the adverse effect of UPF on CD risk.

## Methods

### Study participants

The UKB is an ongoing national prospective cohort project that enrolled over 500,000 volunteers from 22 assessment centers in the UK between 2006 and 2010^6^. UKB collected dietary data with repeated 24-hour diet recalls from 2009-2012 and performed metabolomics profiling at both recruitment (2006-2010) and the first repeated assessment (2012-2013). In the present study, we included participants with at least one available 24-hour dietary recall and excluded: (1) participants reporting extreme energy intake (defined as <800 or >4200 kcal/days for males, <600 or >3500 kcal/days for females)^7^ or a non-typical diet; (2) participants with an IBD diagnosis before recruited. We constructed a circulating metabolite signature of UPF intake in the discovery cohort using individuals with available metabolomic data from the second assessment (n=10,229), where metabolites were measured after the dietary UPF assessment to help mitigate reverse causation. Metabolomic measurements were obtained after dietary assessment, thereby minimizing the potential for reverse causation. The internal validation cohort of the UKB (n=91,306) was leveraged from participants with metabolomic data at recruitment, excluding participants in the discovery cohort. The North West–Haydock Research Ethics Committee granted ethical approval to use the UK Biobank database (REC reference: 21/NW/0157).

One validation cohort is the WHII study^8^. The WHII is a prospective cohort study that recruited 10,308 individuals from the Whitehall departments in London between 1985 and 1988 (Initial phase)^8^. The participants’ dietary information was collected in 1991-1993 (Phase 3) and 1997-1999 (Phase 5), and NMR metabolomic profiles were analyzed through serum samples collected in Phase 5. We included 7,893 WHII participants with available metabolomic data and valid dietary data in the two phases. Given that the WHII study does not collect information on the diagnosis of IBD, we used this cohort to externally validate the metabolic signature of UPF intake identified in UKB. Written informed consent was obtained from all participants at each phase, and the study was approved by the University College London Hospital Committee on the Ethics of Human Research (reference number: 85/0938).

Another validation cohort is the Omics, Nutrition, and Environment Synthesis for IBD (ONE-IBD) Study. The ONE-IBD study is a multicenter cohort study in China that included IBD participants (n=392, 82.1% with CD) and healthy controls (n=360) with available dietary and metabolomic data, who were consecutively recruited between May 2023 and Feb 2025. (**Figure 1**). Dietary information and blood samples used for metabolomics profiling were collected at recruitment. The ONE-IBD study was registered in the UK’s Clinical Study Registry (ISRCTN38557825) and was approved by the Ethical Committee of The Third Xiangya Hospital, Central South University in China (reference number: 25069).

**Figure 1.**
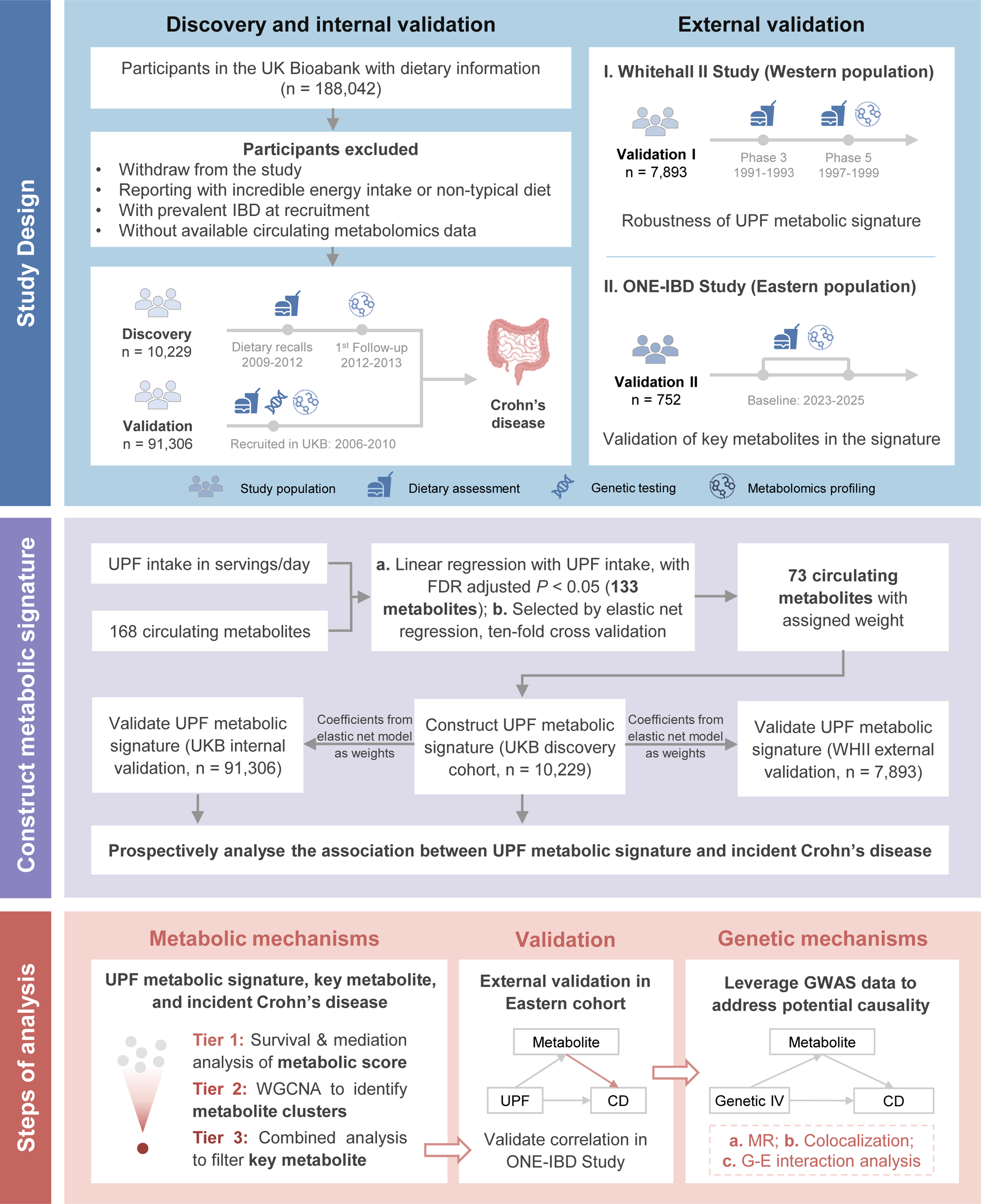
Study flowchart. This study aims to perform a stepwise analysis on the association between UPF, its metabolic signature, and incident CD, and further localize key metabolites and their metabolic and genetic mechanisms in the UPF-CD association. CD, Crohn’s disease; FDR, false discovery rate; G-E interaction, Gene-environment interaction; MR, Mendelian randomization; UPF, ultra-processed food; WGCNA, weighted co-expression network analysis; WHII, Whitehall II Study.

### Dietary assessment and ultra-processed food intake

Dietary information in UKB was collected by a 24-hour Web-based dietary questionnaire (WebQ). Participants were presented with a list of 206 foods and 32 beverages commonly consumed in the UK and selected the number of portions consumed for each food and beverage. The WHII collected dietary data using extensively validated food frequency questionnaires (FFQs) consisting of 127 items. For the dietary information in WHII, we converted the reported food frequency into daily average servings (e.g., 5-6 times a day=5.5 servings/day). Then we assigned the dietary record with the portion size recorded in grams of each food per estimation by the Food Standards Agency^9^. Energy and nutrient content of food items were obtained using *UK McCance and Widdowson’s The Composition of Foods, 6^th^ edition*^10^. In ONE-IBD, a participant’s dietary intake was assessed through an interview-based, photograph-supporting FFQ tailored for the Chinese population, and the energy and nutrients from food were estimated referencing the *China Food Composition Table, 3^rd^ edition*. The UPF was defined by the NOVA classification, and we selected food items used to estimate UPF intake (**Table S1**) based on previous research^4^ ^11^.

### Ascertainment of incident IBD

The primary outcome of interest was incident IBD during follow-up, ascertained through linked hospital inpatient data records (International Classification of Disease Ninth and Tenth Editions [ICD-9 and ICD-10]), coded primary care data, and the death registry. Incident IBD cases were defined as CD (ICD-9 codes 555, ICD-10 codes K50) or UC (ICD-9 codes 556, ICD-10 codes K51) and Read v2/v3 codes mapping to ICD codes (**Table S2**). Previous validation studies in the UK have demonstrated acceptable diagnostic validity of IBD codes in hospital inpatient data, with a reported positive predictive value (PPV) of approximately 87%, and 92% sensitivity in primary care records^12, 13^. The accuracy of the code would increase alongside advances in diagnostic techniques with time^14^. To ensure adequate diagnostic specificity, the primary analysis used a stricter case definition that required at least two recorded IBD diagnostic codes or one code plus at least one record of IBD-related medication use, reducing potential misclassification between CD and UC while maintaining reasonable sensitivity^15^. Participants were followed up from the completed date of the last available 24-hour WebQ to the date of IBD diagnosis, death, loss, or the end of follow-up (England: 31-Oct-2022; Scotland: 31-Mar-2022, and Wales: 31-May-2022), whichever came first.

In the ONE-IBD study, a prospective, clinical-based cohort, the diagnosis of IBD was confirmed by certified gastroenterologists upon participant’s on-site recruitment. According to Chinese clinical practice guideline on the management of Crohn′s disease and ulcerative colitis^16 17^, the diagnosis of IBD at baseline was based on comprehensive review of clinical presentation, endoscopy, radiological, histopathology, and surgical findings. Following enrollment, participants underwent structured longitudinal follow-up at regular intervals around three to six months. Individuals with uncertain diagnoses at baseline, or whose diagnosis was subsequently revised to a non-IBD condition during follow-up, were systematically excluded.

### Construction of a UPF metabolic signature

We used the NMR-measured metabolomics for construction and validation of the UPF metabolic signature in the UKB and validation in WHII. The NMR-based platform provided highly standardized, reproducible metabolic measurements suitable for large population-based cohorts, enabling discovery, validation, and risk prediction with minimal batch effects. Another metabolic profiling methods by ultra-performance liquid chromatography coupled with targeted tandem mass spectrometry (UPLC-MS/MS) with higher sensitivity was used for the Eastern validation cohort ONE-IBD. Details of metabolic profiling are shown in the **Supplementary methods**. The consistency between all metabolites in the metabolomics measurements at both recruitment and the first round of follow-up was displayed in **Figure S1**.

First, we regressed each metabolite on UPF in the multivariable linear model, adjusting for explanatory variables including age, sex, Townsend deprivation index (TDI), physical activity, smoking status, body mass index (BMI), total sugar intake^18^, and total energy intake. The definition and missing rate of these covariates are shown in **Table S3-S4**. The statistical significance of filtered metabolites was evaluated by the Benjamini-Hochberg correction, with an FDR-adjusted P-value<0.05 considered statistically significant.

Second, we applied elastic net regression to filter the metabolites most strongly associated with UPF intake. After elastic net regression, the metabolites that passed the model were further used to construct the metabolic signature score of UPF. The weights of the elastic net model were used to construct the UPF metabolic signature, derived based on the discovery cohort, and we further applied the same weights in the validation cohorts. The UPF metabolic signature score was calculated as a weighted sum of metabolites, obtained by multiplying each metabolite’s standardized concentrations by its corresponding coefficient derived from the elastic net model and then summing these terms across all filtered metabolites.

We further calculated the Spearman correlation coefficient between UPF intake and its metabolic signature in the discovery cohort and in the cohorts for internal and external validation, respectively. To calculate the explained variance (R^2^) of the UPF metabolic signature, we regressed the signature on UPF intake, adjusting for covariates.

### UPF metabolic signature and the incidence of CD

Associations between UPF intake, UPF metabolic signature, and incident IBD, CD, and UC were assessed by Cox proportional hazards regression models, presented as hazard ratios (HRs) and 95% confidence intervals (95% CIs), respectively. Schoenfeld residuals were employed to test the proportional hazards assumption, and no violation was found. We further fitted restricted cubic spline (RCS) models, placing knots at 10th, 50th, and 90th percentiles of UPF intake or UPF metabolic signature, and tested for potential non-linearity by Wald’s test. The mediating proportion was calculated by determining the ratio of the indirect effect to the total effect. We further tested whether the coexistence of genetic susceptibility and UPF intake or its metabolic signature could enhance their overall impact on CD development. This impact was assessed by the additive interaction of the polygenetic risk score (PRS) of IBD or CD and UPF intake or its metabolic signature^19^. In additive interaction, we tested the relative excess risk due to interaction (RERI), the attributable proportion due to interaction, and the synergy index, to evaluate whether UPF intake or its metabolic signature and genetic risk interact additively to increase the risk of IBD. Sensitivity analyses for the survival analysis were further performed to assess associations between UPF intake and its metabolic signature and incident IBD: (1) additionally adjusted for CRP or INFLA-score to test the impact of intrinsic inflammatory level; (2) further excluded IBD or CD cases that occurred within the first two years of follow-up to reduce the reverse-causation bias.

### Identification of key metabolites

Key metabolites were prioritized by integrating multiple complementary analytic approaches, including multivariable linear regression, elastic net model, metabolite clustering by weighted co-expression network analysis (WGCNA), survival analysis, and mediation analysis in the association between UPF metabolic score and incident CD. Details of the WGCNA clustering procedure are displayed in **Supplementary Methods**. The filtering criteria of these analyses include: (1) the top 10 metabolites with the highest absolute β estimates from linear regression; (2) the top 10 metabolites with the highest absolute weights in the elastic net model; (3) metabolites within the filtered cluster in WGCNA; (4) metabolites significantly associated with incident CD in survival analyses; and (5) metabolites showing significant mediation of the UPF-CD association. Metabolites meeting any of these criteria were pooled, and those recurring across approaches were designated as key metabolites.

### External validation in an Eastern cohort

To validate the generalizability of key metabolites in different populations, we leveraged a Chinese cohort with metabolomics data^20^. Details of metabolome profiling are presented in **Supplementary Methods**. In the ONE-IBD validation cohort, we performed a case-control study to assess: (1) the correlation between UPF intake and circulating metabolite levels in IBD, CD and healthy controls; (2) the correlation between circulating metabolite levels and indices reflecting disease prognosis in CD participants, including C-reactive protein (CRP), Crohn’s disease activity index (CDAI), and biological relapse (defined as CRP>10 mg/L during the follow-up); (3) the odds ratio (OR) for CD incidence across different levels of circulating metabolites, in a sub-cohort of newly-diagnosed patients with age- and sex-matched healthy controls.

### Leveraging global GWAS data for causal inference

After pinpointing the key metabolites and validating their mediating effect in ONE-IBD cohort, we leveraged relevant genetic variants from global genome-wide association studies (GWAS) on key metabolites (as exposure) and IBD or CD (outcome) to investigate the potential causality of key metabolites on CD development. For metabolites as an exposure, we extracted summary-level data of published GWAS from 33 cohorts worldwide (distinct from UKB and WHII) that employed the same NMR metabolomics platform, and independent single-nucleotide polymorphisms (SNPs) associated with metabolites were subsequently selected as instrumental variables^21^. The genetic instrumental variables selected were displayed in **Table S5**. For IBD and CD as outcomes, we leveraged GWAS data from UKB, the FinnGen study, and the International Inflammatory Bowel Disease Genetics Consortium (IIBDGC)^22-24^. After extracting SNPs associated with IBD and CD, we conducted a meta-analysis of the three GWAS to obtain pooled genetic variants of each outcome. MR analysis was used to assess the link between genetically predicted circulating metabolite levels and incident IBD or CD, based on the results of epidemiological analysis^25^. A *P*<5×10^-8^ was considered as genome-wide significant, and the inverse variance weighted method was applied^26^. As sensitivity analyses for MR, we further employed the weighted median, MR Egger, MR Pleiotropy residual sum and outlier approach (MR-PRESSO), and leave-one-out MR analysis to validate the potential causality between key metabolites and incident IBD. Colocalization analyses with single traits (‘coloc’ method) and multi-traits (‘HyPrColoc’ algorithm) were performed to identify shared causal variants between key metabolites and incident IBD or CD^27 28^. Pair-Wise Conditional analysis and Colocalization analysis (PWCoCo) were conducted to scan non-primary signals in the region which colocalize^29^. Gene-environment (G-E) interaction analysis was performed in the UKB to validate whether genotypes of the filtered genetic loci could influence the association between UPF intake, UPF metabolic signature, and incident IBD or CD^30^. Details of the acquisition and processing of GWAS data, MR analysis, colocalization, and G-E analysis were presented in **Supplementary Methods**.

All analyses were performed using R software, version 4.2.1. A two-sided *P* < 0.05 was considered significant.

### Patient and public involvement

Patients and/or members of the public were not involved in the design, conduct, reporting, or dissemination plans of this research.

## Results

### Characteristics of study participants

The study included 10,229 participants in the discovery cohort and 91,306 participants from the UK Biobank (UKB) in the internal validation cohort. External validation was performed in two independent cohorts: the Whitehall II (WHII) study (n=7,893) and the the Omics, Nutrition, and Environment Synthesis for IBD (ONE-IBD) Study (n=752). Baseline characteristics across cohorts are presented in **Table S6**. Compared to UKB, participants from the WHII study included more men and were overall younger, with higher socioeconomic levels (reflected by higher TDI), lower BMI, higher total energy intake, and higher UPF intake in servings, grams, and energy proportion per day. Participants in the Eastern cohort ONE-IBD were younger and had a lower BMI compared with the Western cohorts.

### Metabolic signature of UPF intake

Out of all 168 serum metabolites, 133 metabolites were significantly associated with UPF intake (false discovery rate [FDR]-adjusted *P*<0.05) in linear regression analysis. Subsequent elastic net regression identified a metabolic signature consisting of 73 metabolites. The metabolic signature indicates a shift toward more unsaturated fatty acid traits, enriched with DHA, a remodeled energy metabolism profile marked by higher glucose levels with pyruvate accumulation and reduced reliance on acetate-derived acetyl-CoA input, and inflammatory-related metabolic perturbations, accompanied by changes in albumin and creatinine. These metabolites comprised 26 sterol lipids, 15 lipoprotein particles, 10 glycerophospholipids, 8 glycerolipids, 6 fatty acids, 3 carbohydrates and their derivatives, 3 metabolites related to fluid balance and inflammation, and 2 ketone bodies, with most of them being lipid-related metabolites (**Table S7**). The metabolic signature exhibited a significant correlation with UPF intake across the discovery cohort and the two validation cohorts, with Spearman correlation coefficients ranging from 0.20 to 0.25, showing relatively high correlation compared with previous study in diet-related metabolomic signature^31^ (**Figure 2A-C**). Metabolites selected and their weights derived from the elastic net model ranged from -1.28 to 1.52 (**Figure 2D**).

**Figure 2.**
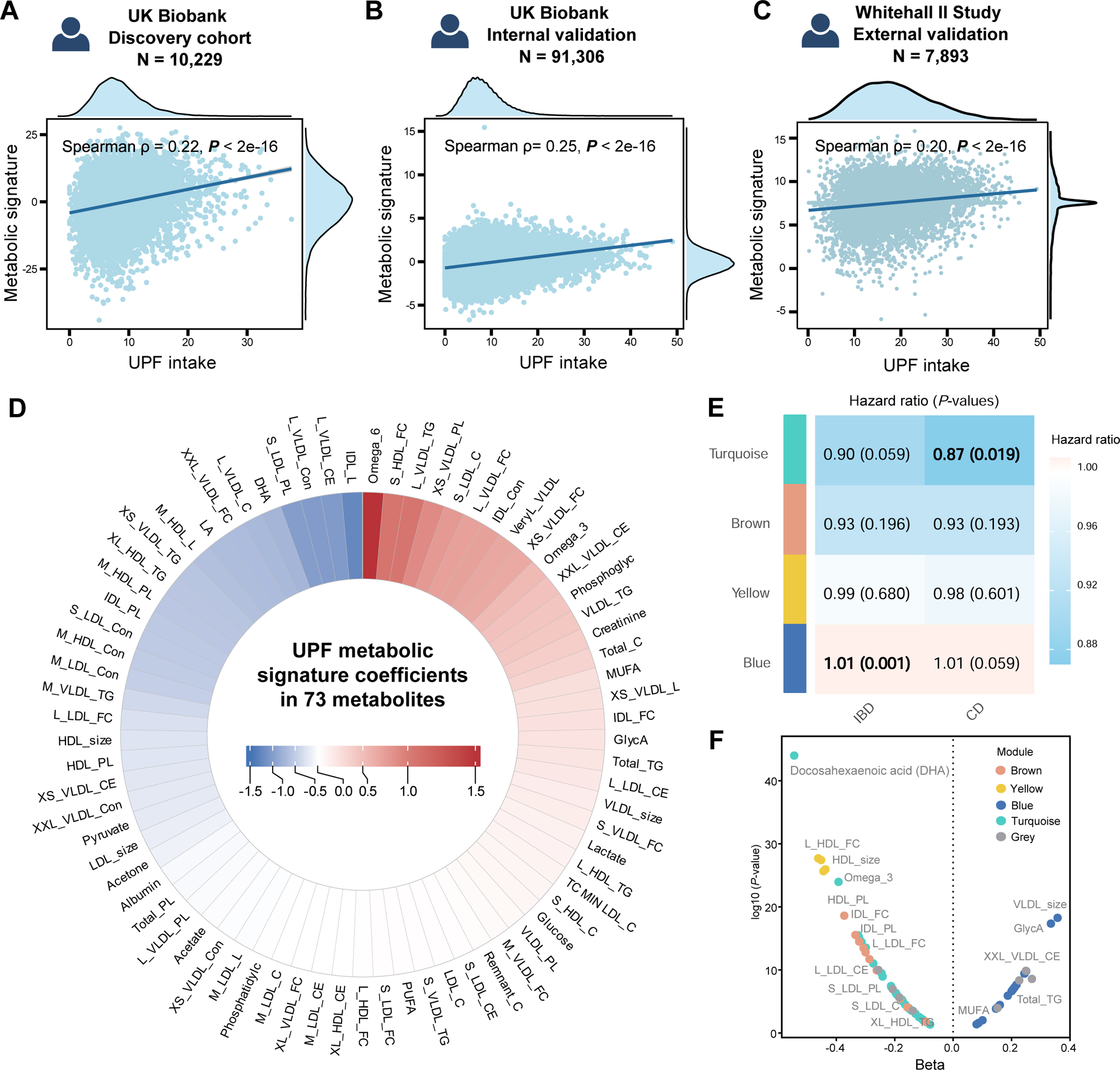
The metabolic signature of UPF intake. Spearman correlation coefficients between UPF metabolic signature and UPF intake in the UK Biobank (A-B) and WHII study (C). Seventy-three metabolites passed the elastic net model with weights used to construct the UPF metabolic signature (D). The module-trait association between WGCNA metabolite modules and incident IBD, CD, and UC; the values and color of the square refer to the Pearson correlation coefficient, with the values in the bracelets refer to the *P*-values (E).The beta values in the linear regression, displaying 73 metabolites filtered by elastic net model on UPF intake, with different colors of dots refers to modules categorized by WGCNA (F). CD, Crohn’s disease; DHA, docosahexaenoic acid; IBD, inflammatory bowel disease; UC, ulcerative colitis; UPF, ultra-processed food; WGCNA, weighted co-expression network analysis.

### UPF intake and its metabolic signature associated with IBD incidence

During a median follow-up of 10.9 years, we documented 295 CD cases and 684 UC cases among 188,042 participants with available UPF intake data. UPF intake was positively associated with increased risk of IBD. Individuals in the highest tertile of UPF serving intake presented an elevated IBD risk compared to those in the lowest tertile (HR _T3 vs T1_=1.21, 95% CI 1.03-1.43, *P*=0.020). This positive association was stronger in CD (HR _T3 vs T1_=1.52, 95% CI 1.11-2.06, *P*=0.008), whereas the association with UC did not reach statistical significance (**Figure 3 A-B, Table S8**). Given that the associations were consistently stronger and statistically significant for CD, whereas no significant associations were observed for UC, subsequent analyses were therefore focused on CD. The metabolic signature of UPF intake showed a significant association in IBD, especially in CD. A higher metabolic signature indicated a higher risk of IBD in both the discovery (HR_high vs low_=2.20, 95% CI 1.09-4.45, *P*=0.006) and validation cohort in UKB (HR_T3 vs T1_=1.65, 95% CI 1.25-2.18, *P*<0.001). The UPF-related metabolic signature was significantly associated with increased risk of both CD but not UC across cohorts (**Figure 3A-B, Table S8**). Restricted cubic spline (RCS) presented no non-linear association between both UPF intake and its metabolic signature and incident CD and IBD (*P*-nonlinearity>0.05, **Figure 3C-D**, **Figure S2**). In mediation analysis, the UPF metabolic signature mediated 21.7% (*P*<2e^-16^) of the association between UPF intake and incident CD (**Figure 4A**).

**Figure 3.**
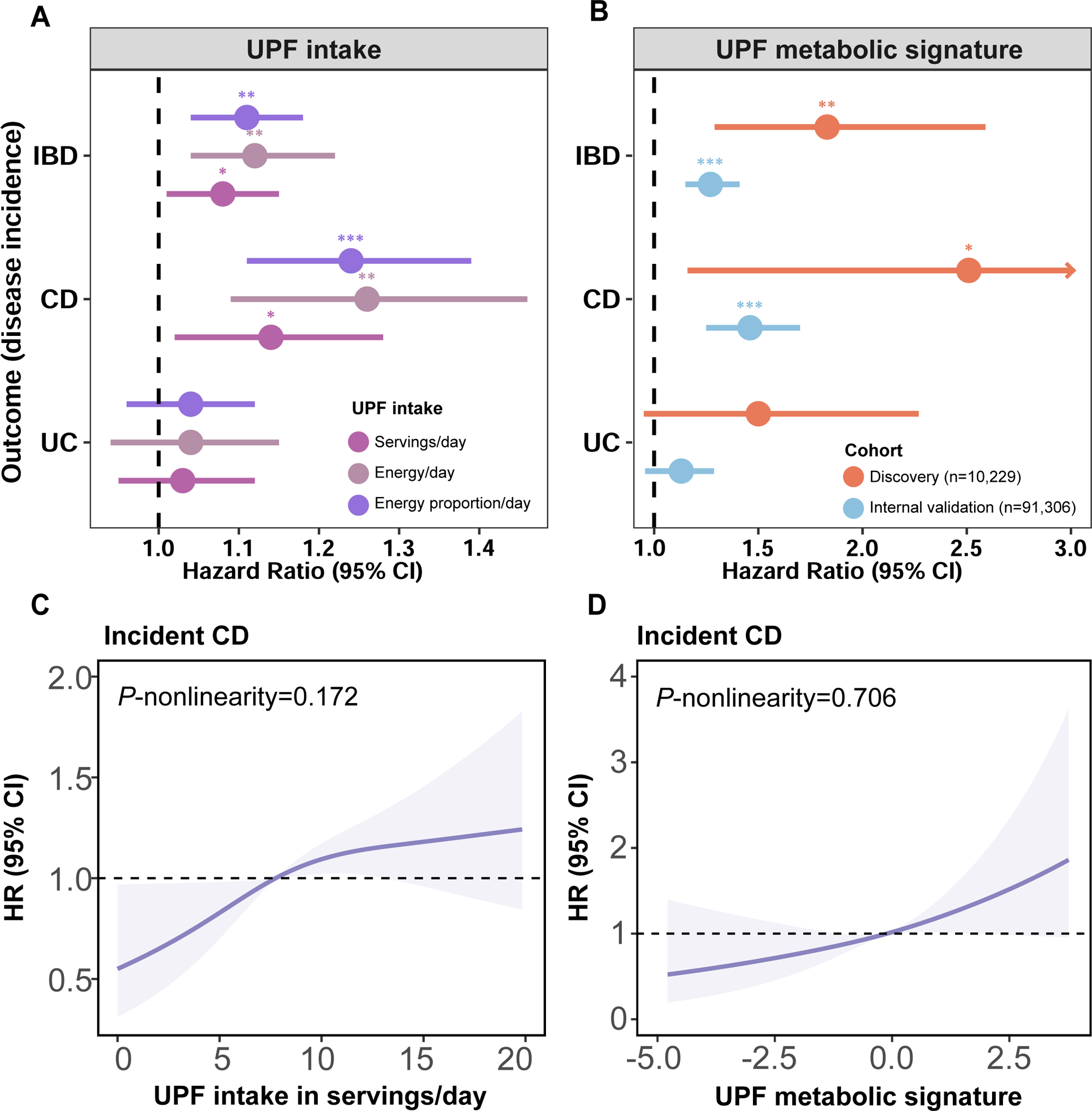
Survival analysis of the UPF intake and incident CD, and the gene-diet interaction. The association between UPF intake, UPF metabolic signature, and the risk of IBD, CD, and UC (A-B). The dose-response association between UPF intake, UPF metabolic score, and incident CD (C-D). CI, confidence interval; HR, hazard ratio; IBD, inflammatory bowel disease; UPF, ultra-processed food.

**Figure 4.**
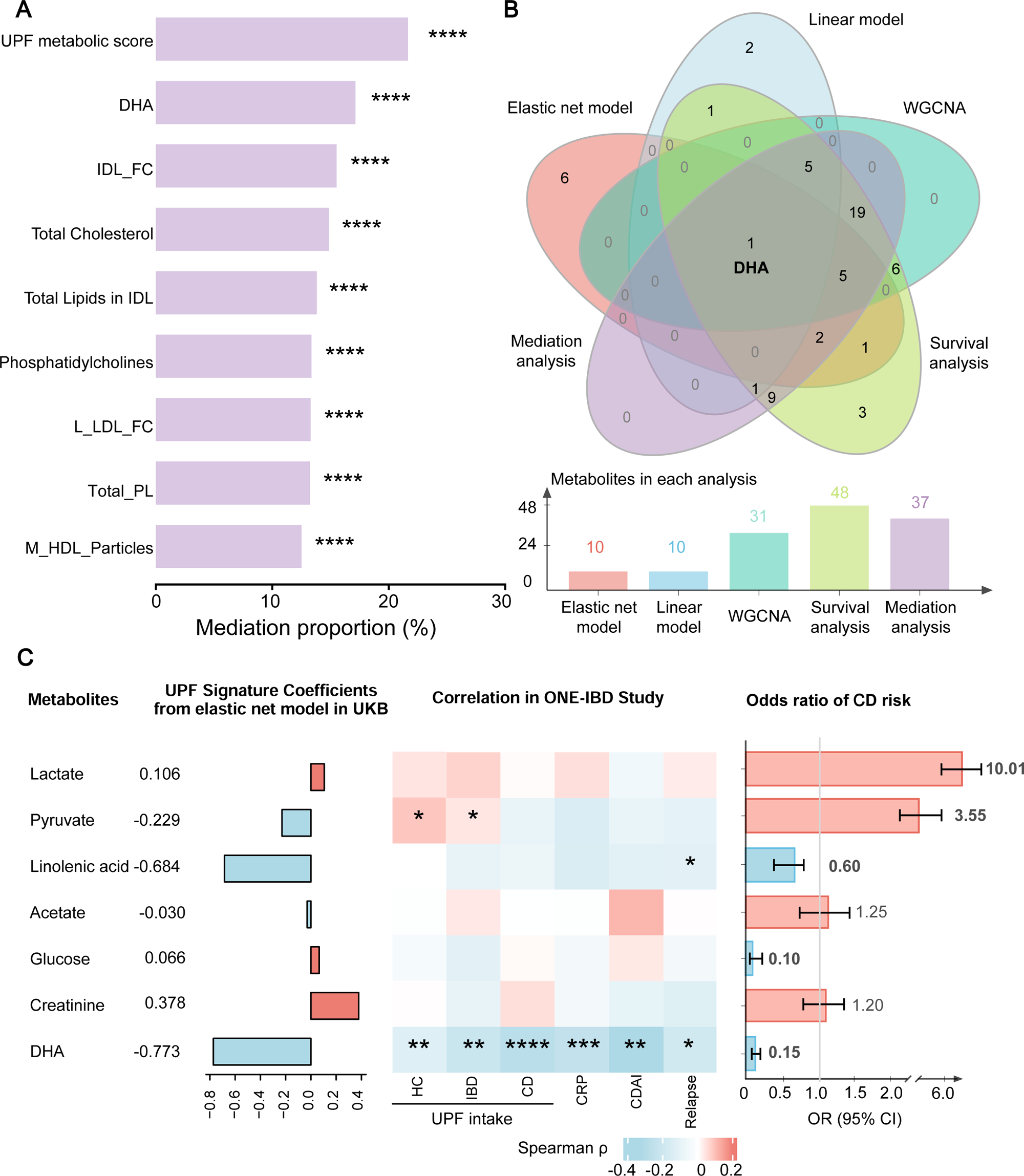
Analysis and validation of DHA as the key metabolite mediating the association between UPF and incident CD. The mediation proportion of UPF metabolic score and key metabolites with the highest mediation proportion (A). DHA was pinpointed in the UPF-CD association as the only one overlap metabolite filtered through five distinct analyses, including the top ten metabolites with the highest weight (absolute values) in the linear model and elastic net model, the metabolites in the turquoise module derived from WGCNA, the metabolites presenting significant association with incident CD in the survival analysis, and the metabolites with the highest mediation proportion in mediation analysis (B); association between key metabolites, UPF intake, and CD disease prognostic indices in ONE-IBD validation cohort (C). CD, Crohn’s disease; CDAI, Crohn’s disease activity index; CRP, C-reactive protein; DHA, docosahexaenoic acid; HC, healthy control; IBD, inflammatory bowel disease; UPF, ultra-processed food; WGCNA, weighted co-expression network analysis. \**P*<0.05, \*\**P*<0.01, \*\*\**P*<0.001.

### Interaction with the genetic susceptibility of CD

Regarding the genetic risk of CD, participants with high UPF intake and high genetic risk presented the highest risk for developing CD compared to those with low UPF intake and low genetic risk (HR =2.06, 95% CI 1.46-2.92, *P*<0.001). We observed evidence of an additive interaction between UPF intake and incident CD (relative excess risk due to interaction [RERI]=0.62; 95% CI 0.02-1.22, *P*=0.021). Correspondingly, participants with high metabolic signature score and high genetic susceptibility presented increased CD risk compared to those with both features at the low level (HR=2.07, 95% CI 1.23-3.47, *P*=0.006). The additive interaction (*P* for RERI=0.007) was significant for the analysis of UPF metabolic signature (**Table S9**). RCS displayed linear associations when stratified by genetic risk, and the risky effect of UPF on incident CD was stronger in individuals with higher genetic susceptibility (**Figure S3**).

### Metabolites clustered by WGCNA

We further explored the metabolic mechanisms through clustering metabolites in IBD, including CD. The WGCNA framework was applied to cluster the 73 metabolites filtered by elastic net regression. Four metabolite clusters were derived from the WGCNA network (presented as modules in blue, turquoise, yellow, and brown), plus one module (in grey) representing unclassified features. The four identified modules contain 65 metabolites ranging in size from 4 to 31 members (**Table S10**). The module showing strongest protective effect against incident IBD (in turquoise in **Figure 2E**) was rich in lipids, including sterol lipids, fatty acids, and glycerophospholipids, while the module with the strongest hazardous effect (in blue) was particularly rich in lipids and lipoproteins. When combining the z-score, only the module with the strongest IBD-protective effect showed a significant association with incident CD (HR_turquoise_=0.87, 95% CI 0.77-0.98, *P*=0.019, **Table S11**). The correlations between the module eigengenes and incident IBD and CD are displayed in **Figure 2E** and **Table S12**.

### Identifying DHA as the key metabolite

Based on the identified metabolite modules, we selected the metabolites in the turquoise module for single-metabolite analyses to identify key metabolites. Through mediation analysis, we identified 37 metabolites with significant mediation effects (**Table S13**), among which DHA presented the strongest mediating proportion (17.1%, **Figure 4A**). In survival analysis, we identified 36 metabolites significantly associated with incident IBD (48 metabolites with incident CD, **Table S14**). We also listed metabolites clustered by WGCNA regressed on UPF intake by linear model, within which DHA showed the strongest effect with negative weight (β=-0.544, **Figure 2F**). Combining results from linear model, elastic net model, survival analysis, mediation analysis, and the turquoise module characterized by WGCNA, DHA stood out as the only metabolite that performed significantly in all analyses (**Figure 4B**). In sensitivity analyses, we observed a consistent association between UPF metabolic signature and incident CD, after further adjusting for CRP or INFLA-score, or excluding incident cases in the first two years of follow-up (**Table S15-S16**).

In the Eastern validation cohort ONE-IBD, compared to 73 metabolites used to construct UPF metabolic signature, we identified seven metabolites that were tested in metabolomics data. DHA was inversely correlated with UPF intake in both healthy controls (Spearman ρ=-0.16) and CD participants (ρ=-0.28). DHA was also correlated with lower disease activity, evidenced by lower levels of CRP and CDAI, and lower rate of biological relapse (**Figure 4**, **Table S17**). Furthermore, circulating DHA levels were inversely associated with CD risk in the case-control study, showing a 79% reduction in CD risk per standard deviation increase in circulating DHA (OR=0.21, 95% CI 0.16-0.28, *P*<0.001, **Table S18**).

MR analysis further elucidated the potential causal association between DHA and CD. The genetically predicted circulating levels of DHA showed a significant inverse association with IBD and CD (**Table S19**). For DHA, the ORs of IBD and CD were 0.79 (95% CI 0.72-0.86, *P*=2.87e^-07^) and 0.73 (95% CI 0.63-0.84, *P*=2.11e^-05^) using the IVW method. These results were consistent when applying the weighted median, MR Egger, MR-PRESSO, and leave-one-out MR analyses (**Table S20, Figure 5A-C**). In the colocalization analysis, we identified rs174546 in *FADS1* as a key gene locus for sharing a causal variant between circulating DHA levels and the risk of CD (**Figure 5D**), with a relatively high PPH4 in CD (95.8%) using ‘coloc’ analysis, and further validated by HyPrColoc algorithm, the multi-trait PPFC (PPE) was 73.65% (34.79%) in IBD and CD (**Figure 5E, Figure S4**). PWCOCO analysis did not find colocalized signals other than rs174546 out of the region (**Table S21**). Expression quantitative trait locus (eQTL) analyses indicate that rs174546 influences the expression of *TMEM258, MYRF, FADS2*, *FADS1,* and *ENSGO0000289268* across various tissues (**Figure S5**). The serum DHA level was higher in CC homozygotes of rs174546 compared to T allele carriers (**Figure 5F**). In the G-E interaction analysis, the positive association between UPF intake and incident CD was amplified in rs174546 T-allele carriers, while the association for the UPF metabolic signature was attenuated in the same group (**Figure 5G**).

**Figure 5.**
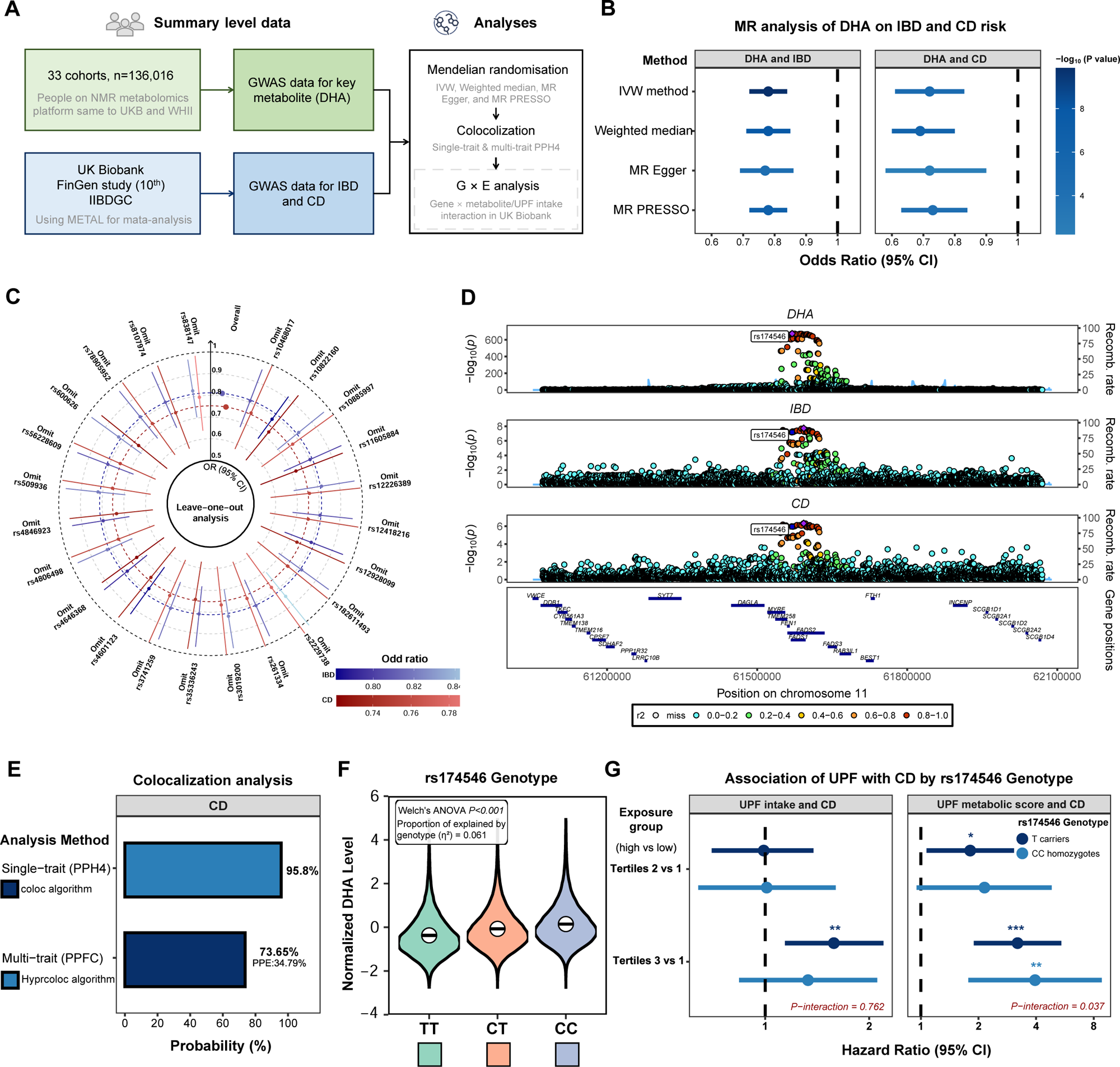
Mendelian Randomization and colocalization analysis of DHA and incident CD, validated by G-E interaction analysis. Study flowchart of the analyses (A); Mendelian Randomization analysis of circulating DHA level on IBD and CD risk using different methods (B); Leave-one-out MR sensitivity analysis of DHA on IBD and CD risk (C); colocalization analysis using HyPrColoc to identify key genetic locus (D); results of the colocalization analysis applying two methods, indicating evidence for shared causal variation between circulating DHA levels and incident CD (E); DHA levels among varied genotypes of the SNP rs174546 (F); association of UPF intake, UPF metabolic score with CD risk stratified by rs174546 genotype (G). CD, Crohn’s disease; DHA, docosahexaenoic acid; GWAS, genome-wide association study; IBD, inflammatory bowel disease; IIBDGC, International Inflammatory Bowel Disease Genetics Consortium; PPFC, posterior probability of full colocalization; PPE, proportion of PPFC explained by the listed SNP; PPH4, posterior probability of hypothesis 4; SNP, single nucleotide polymorphism.

## Discussion

Leveraging large-scale prospective cohorts and multi-omics profiling, we developed and validated a circulating metabolic signature that captures UPF intake and predicts future CD risk, with DHA being the key metabolic mediator that accounts for 17.1% of the UPF-CD association. Rather than being an additive risk factor, our findings suggest that UPF intake contributes to CD risk through a relative deficiency of DHA and other protective metabolites, leading to a disrupted metabolic profile. This signature, characterized by low DHA levels, an imbalance in unsaturated fatty acids, and a remodeled energy metabolism profile, provides a practical metric to identify individuals at elevated CD risk and to stratify susceptibility based on metabolome-defined heterogeneity in dietary responses to UPF. Moreover, our findings support gene-diet interplay, with DHA-related mechanisms being more pronounced among individuals with higher genetic risk and specific *FADS1* genotypes.

Accumulating epidemiological evidence supports the association between higher UPF consumption and the increased risk of CD^4 32 33^, while the identification of its biological mechanisms is still in an early stage. Circulating metabolites, as indicators of dietary response, can serve as the bridge between dietary intake and the subsequent health effects^34^. A narrative review summarized the contribution of UPF in CD pathogenesis, pointing out the adverse impact of UPF on gut microbial dysbiosis and damaged mucosal barriers, mediated by circulating fatty acids^2^. The processing byproducts of UPF, like glycation end products and polycyclic aromatic hydrocarbons (PAHs), can reduce beneficial bacteria that produce short-chain fatty acids, the lack of which can subsequently exacerbate lipid-induced inflammation^35^. Additionally, UPF displaces fiber-rich foods and leads to nutritional deficits that negatively affect mucosal immunity, with underlying mechanisms linked to excessive oxidative stress and disrupted lipid metabolism in the gut^36^.

Circulating plasma metabolome results from the synergistic effect of metabolism-modulating factors such as lifestyle and diet and reflects the overall homeostasis of the internal environment^37^. Recent clinical studies have identified several metabolite indicators associated with UPF consumption and their adverse effects on multiple diseases. Regarding UPF-related metabolic traits, a cohort study involving over 3,000 participants in the United States found a metabolic signature significantly associated with UPF intake, indicating UPF-related metabolite markers can be applied in prediction of chronic kidney disease^5^. For individuals who underwent metabolome testing in the UKB, a prospective cohort study identified a metabolomic signature comprising 34 metabolites related to UPF intake that was significantly associated with risk of MASLD^38^. Regarding the component of UPF, another study constructed a metabolomic score related to processed meat consumption, and found that processed meat-related metabolomic signatures are associated with elevated risk of ischemic heart disease^39^. These studies corroborated our hypothesis and support an association between UPF intake, its related metabolic profiles, and multiple diseases. Our study, in particular, observed the key role of UPF-related metabolic signatures in predicting the risk of CD.

Notably, we identified DHA as the critical circulating metabolite in the UPF metabolic signature, specifically mediating the UPF-CD association. DHA is a beneficial n-3 polyunsaturated fatty acid (PUFA) rich in fresh, natural foods like fish (e.g., salmon and tuna), shellfish, algae, and egg yolks^40^. It is an essential nutrient with anti-inflammatory effects that facilitates brain development, cardiovascular function, and immune regulation^41^. Increased consumption of UPF may diminish circulating DHA concentration. A population-based study found that participants with higher frequency of UPF intake show lower intakes of DHA-rich foods (e.g., fish, soybean products), and lower levels of circulating DHA^42^. A biological explanation might be that high content of unhealthy fats rich in UPF may interfere with the balance of n-3 PUFA and disrupt gut microbiota fermentation, further contributing to the decline in circulating DHA levels^43^. In previous studies, DHA is the common metabolic indicator of UPF intake. A cross-sectional study revealed that UPF consumption was negatively associated with circulating PUFAs (including DHA and conjugated linoleic acid) and other 114 metabolic traits in British children^44^. Studies regarding the protective effect of DHA on CD primarily focused on microbiome alteration and immune function restoration, which echoed the disruption mechanisms of excessive UPF consumption. Research in mice has found that DHA may alleviate the gut inflammation and block colitis by enhancing the colonic mucus barrier^45^.

In line with previous literature, maintaining a DHA-rich dietary pattern has been included as a key component of healthy lifestyle recommendations for reducing CD risk^46^. Our findings extend this perspective by suggesting that the adverse effects of ultra-processed food intake may arise not solely from harmful additives, but from a relative depletion of protective nutrients such as DHA. Dietary supplementation of DHA has been linked to restoration of gut dysbiosis, with lowered serum triglyceride levels and better prognosis in individuals with diabetes mellitus^47^. Considering the interaction between DHA and other dietary components, research found that DHA-enriched phosphatidylcholine (DHA-PC) can ameliorate the high-fat diet-induced MAFLD through regulating circulating fatty acids, indicating DHA as a dietary intervention target for gastrointestinal diseases^48^. Furthermore, DHA can restore the intestinal barrier function through pathways related to PPARγ and show protective effects against the gut damage caused by chemicals^49^. One animal study found that DHA alleviates inflammation by reducing colon epithelium shredding and improving mucosal structure, offering a therapeutic strategy to mitigate colitis-induced effects and manage CD^50^.

The external validation in the independent ONE-IBD cohort, an East Asian population, provides evidence for the generalizability of our findings. The consistent presence of inverse associations between DHA and UPF intake in both healthy controls and newly-diagnosed CD participants suggests that the deleterious metabolic consequence of UPF consumption, namely, the depletion of circulating DHA, is observable despite differences in dietary cultures and genetic backgrounds. More importantly, the protective association between higher DHA levels and reduced CD risk observed in the ONE-IBD cohort (**Table S19**) reinforces the main finding from our UKB analyses. This suggests that the protective role of DHA against CD may be mediated by biological mechanisms shared across populations. The inverse correlations observed between circulating DHA levels and multiple clinical disease activity indices (CRP and CDAI) among IBD patients, furthermore, have clinical implications. These findings indicated that DHA may not only influence disease susceptibility but also potentially modify disease behavior after onset^51^. Collectively, these validation results solidify DHA as a key, relevant metabolic entity in the UPF-CD association, underscoring its potential for CD prevention in both Western and Eastern populations.

Results from MR and colocalization analysis further support a potentially causal role of circulating DHA in incident CD. In colocalization, rs174546 at the *FADS1* locus emerged as a shared genetic signal linking DHA levels and IBD risk, with high posterior probability of a common causal variant (PPH4: 95.8% for CD). The rs174546 is located in the 3’ untranslated region (3’UTR) variant of *FADS1* (chromosome 11q12.2), a locus implicated in regulating FADS1/Δ5-desaturase activity and long-chain PUFA biosynthesis, including DHA^52^. The variation of rs174546 from C to T was found to lower DHA levels and its proportion to total fatty acids; this variation may diminish the protective role of circulating DHA on counteracting the UPF’s effect in the CD pathogenesis^53^. Prior studies have linked the T allele to reduced Δ5-desaturase activity, resulting in lower circulating DHA levels and a decreased proportion of DHA among total fatty acids, which may weaken the protective role of DHA in counteracting UPF-related pathogenic processes in CD. Therefore, rs174546-T carriers may have a diminished capacity for endogenous DHA formation and be more dependent on exogenous DHA intake^53^. Consistently, our G-E analysis suggests that individuals with genetic make-up suggest lower DHA (rs174546 T allele carriers) may be at greater risk of CD in the context of higher UPF intake. Notably, the relationship between the UPF metabolic score and CD appeared attenuated among those with lower DHA levels, potentially reflecting that DHA is a dominant component embedded within the UPF-related metabolomic profile. Collectively, these findings reinforce DHA as a central metabolic link between UPF intake and CD risk.

To our knowledge, this is the first study to explore the association between the metabolic signature of UPF and CD. The metabolic signature was reproducible in the external WHII study and remained robust across sensitivity analyses. Importantly, our study suggest that UPF-related CD risk may be driven by a relative depletion of protective metabolites, with DHA emerging as a potential intervention target in the UPF-CD association, highlighting its future clinical application to predict and prevent future incident CD linked to increased UPF intake. The all-encompassing data of the UKB, WHII, ONE-IBD study, and the global GWAS data enabled us to strengthen the robustness and generalizability of our findings through cross-cohort replication and genetic triangulation (i.e., integrating genetic evidence by GWAS-based analyses with observational results), facilitating rigorous sensitivity analyses and causal inference. Importantly, validation in the Eastern ONE-IBD cohort extended the applicability of our findings beyond predominantly Western populations. Nevertheless, some limitations warrant discussion. First, a diet high in UPF shares multiple nutrients and food components with other dietary patterns, and the identified metabolic signature may also link with other, non-dietary factors, which may limit the specificity of this metabolic signature reflecting the dietary UPF intake. Therefore, we calculated the correlation coefficient and validated the signature by an external WHII study to assess the stability of the metabolic signature. Second, heterogeneity may exist as UPF intake was estimated using different dietary questionnaires across the UKB, WHII, and ONE-IBD. Nevertheless, despite cohort-specific instruments and scoring approaches, we observed consistent associations across cohorts, supporting the robustness and reliability of our findings. Third, the NMR metabolomics platform applied in UKB and WHII did not encompass all circulating metabolites potentially related to dietary change; which means that inclusion of undetected metabolites in future research could add increased precision to the current signature. Finally, though we have validated the reproducibility of the metabolic signature and the benefit of DHA in different cohorts, the biological mechanisms of our findings still need to be validated by *in vivo* experiments, especially the effect of the *FADS1* gene and the circulating DHA level.

In summary, our study identified a metabolic signature that mediates the association between UPF intake and incident CD. This signature reflects personalized metabolic responses to UPF exposure and is positively associated with future CD risk. Notably, our findings indicate that UPF-associated CD risk may arise not only from harmful dietary additives, but also from a relative reduction in protective metabolites, particularly DHA. Individuals with higher genetic susceptibility to CD appeared more vulnerable to these DHA-related metabolic alterations, with genetic evidence implicating the FADS1 locus.

These findings revealed the potential role of DHA in reversing the detrimental impact of UPF on CD risk, providing evidence on precision nutrition for CD prevention. Critically, the reproducibility of these findings was demonstrated in the Eastern population (ONE-IBD), where DHA consistently exhibited an inverse association with UPF intake and a strong protective effect against CD, supporting its generalizability across different ethnic and dietary backgrounds. These findings underscore identifying and monitoring metabolic status as a more comprehensive approach in reflecting dietary response of UPF, and suggest a potentially effective ingredient in reversing the adverse health effects of an unhealthy diet in the precision prevention of CD.

## Abbreviations

BMI: body mass index
CD: Crohn’s disease
CDAI: Crohn’s disease activity index
CI: confidence interval
CRP: C-reactive protein
DHA: docosahexaenoic acid
FDR: false discovery rate
FFQ: food frequency questionnaire
GWAS: genome-wide association study
HR: hazard ratio
IBD: inflammatory bowel disease
IIBDGC: International Inflammatory Bowel Disease Genetics Consortium
MR: Mendelian Randomization
NMR: nuclear magnetic resonance
OR: odds ratio
PPFC: posterior probability of full colocalization
PPE: proportion of PPFC explained by the listed SNP
PPH4: posterior probability of hypothesis 4
PUFA: polyunsaturated fatty acid
RCS: restricted cubic spline
SNP: single nucleotide polymorphism
TDI: Townsend deprivation index
UC: ulcerative colitis
UKB: UK Biobank
UPF: ultra-processed food
WGCNA: weighted co-expression network analysis

## Data sharing statement

The results are presented in the article and supplemental materials. The UK Biobank is available to researchers with approval (www.ukbiobank.ac.uk/). Whitehall Il data are available to bona fide researchers for research purposes. Please refer to the Whitehall II data sharing policy at the website (www.ucl.ac.uk/whitehalll/data-sharing). Data access requests regarding the ONE-IBD Study should contact the corresponding author. Approval is at the discretion of the principal investigator and the sponsor, contingent upon the proposed research objectives, data availability, and intended data use.

## Acknowledgements

We are much obliged to the administrative team and participants of the UK Biobank (application number: 232231). We thank all participants in the Whitehall Il Study, Whitehall Il researchers and support staff who make the study possible. The UK Medical Research Council (MR/KO13351/1; G0902037), British Heart Foundation (RG/13/2/30098), and the US National Institutes of Health (RO1HL36310, RO1AG013196) have supported collection of data in the Whitehall II Study (application number: 0729). And we thank all participants and researchers of the Omics, Nutrition, and Environment Synthesis for IBD Study (ONE-IBD, registration number: ISRCTN38557825).

## Author contributions

All authors have read and approved the final version of the manuscript. Sidan Wang (Design: equal; Methodology: equal; Visualization: equal; Writing-original draft: leading; Writing-review & editing: equal). Lintao Dan (Methodology: equal; Formal analysis: leading; Visualization: equal; Writing-original draft: equal; Writing-review & editing: equal). Xixian Ruan (Methodology: equal; Formal analysis: equal; Visualization: equal; Writing-original draft: equal; Writing-review & editing: equal). Judith Wellens (Conceptualization: supporting; Methodology: equal; Writing-review & editing: supporting). Yuhao Sun (Methodology: equal; Formal analysis: supporting; Writing-original draft: supporting; Writing-review & editing: equal). Jialu Yao (Methodology: supporting; Writing-review & editing: supporting). Li Tian (Methodology: supporting; Writing-review & editing: supporting). Rahul Kalla (Methodology: supporting; Writing-review & editing: supporting). Evropi Theodoratou (Conceptualization: supporting; Methodology: supporting; Writing-review & editing: supporting). Shuai Yuan (Design: supporting; Methodology: supporting; Writing-review & editing: supporting). Susanna C Larsson (Methodology: supporting; Writing-review & editing: supporting). Jonas F Ludvigsson (Methodology: supporting; Writing-review & editing: supporting). Laurent Peyrin-Biroulet (Design: supporting; Writing-review & editing: supporting). Jack Satsangi (Methodology: supporting; Writing-review & editing: supporting). Fernando Magro (Conceptualization: supporting; Methodology: supporting; Resources: equal; Writing-review & editing: supporting). Xue Li (Conceptualization: equal; Methodology: supporting; Resources: equal; Funding acquisition: equal; Writing-review & editing: supporting). Xiaoyan Wang (Conceptualization: equal; Project administration: equal; Resources: equal; Funding acquisition: equal; Writing-review & editing: supporting). Jie Chen (Conceptualization: leading; Formal analysis: equal; Project administration: leading; Resources: equal; Funding acquisition: equal; Writing-review & editing: equal).

