## Supplementary material for "Integrated metabolomics and genetic analyses reveal loss of protective docosahexaenoic acid as a key driver linking ultra-processed food to Crohn’s disease risk"

### **Supplementary Methods**

***Metabolomics profiling***

Non-fasting Ethylenediaminetetraacetic acid (EDTA) plasma samples were collected and stored at -80°C for UK Biobank (UKB) participants in 2012-2013 (discovery cohort) and 2006-2010 (internal validation cohort), and for participants in the Whitehall II Study (WHII) in 1997-1999, as described in previous literature^1^. Both studies used the Nightingale (Helsinki, Finland) platform, a high-throughput nuclear magnetic resonance (NMR) metabolomics platform for metabolite assays. NMR spectra were automatically quality-checked, with flags applied for potential degradation or quantification issues. Platform reproducibility was further supported by CE marking and ISO 13485 certification, with strong correlations (typically >0.9) observed when compared with conventional clinical biochemistry measurements in external validation cohorts. Batch effects were minimal across consecutive analytical batches, reflecting standardized pre-analytical handling and minimal sample preparation. The metabolomics of UKB analyzed a total of 251 metabolic biomarkers, including 170 absolute concentrations and 81 derived ratio measurements. Similarly, the WHII measured NMR metabolomics, including a total of 233 metabolic biomarkers, and 169 absolute concentrations were included. To ensure accuracy, we finally included 168 metabolites measured as absolute concentrations collected in both UKB and WHII in the procedures to construct the metabolic signature. We further excluded participants with missing metabolomics data in elastic net regression when calculating the weight of each metabolite. Participants with outliers for each metabolite were also excluded from analysis, defined as values more than 9 standard deviations (SDs) away from the mean value.

In the validation cohort, the Omics, Nutrition, and Environment Synthesis for IBD Study (ONE-IBD), blood samples were collected at the time of Food Frequency Questionnaire (FFQ) collection, and stored at -80℃ freezer immediately. Plasma samples were extracted for circulating metabolomics profiling. Targeted metabolomic profiling was performed using ultra-performance liquid chromatography coupled with tandem mass spectrometry (UPLC-MS/MS). Briefly, 20 μL of plasma samples, thawed on ice, were protein-precipitated with 120 μL of ice-cold methanol containing internal standards. After vortexing and centrifugation, the supernatant was subjected to a derivatization reaction at 30°C for 60 minutes. The derivatized samples were then diluted with a 50% methanol solution and centrifuged again before UPLC-MS/MS analysis. Chromatographic separation was achieved on an ACQUITY UPLC BEH C18 column (1.7 μm, 2.1 × 100 mm) maintained at 45°C. Mass spectrometry was conducted in negative ion mode with a capillary voltage of 1.2 kV, a source temperature of 150 °C, and a desolvation temperature of 550 °C. Quality control was obtained by mixing a small aliquot of each biological sample in the study set. Metabolite data were acquired using multiple reaction monitoring (MRM), with cone voltage and collision energy optimized via the QuanOptimize application manager (Waters). Raw UPLC-MS/MS data were processed and quantified using TargetLynx version 4.2 (Waters Corp., Milford, MA, USA). Peak integration, calibration, quantification, quality control, and batch effect correction were performed using TMBQ software (v1.0; Human Metabolomics Institute, Shenzhen, China), in accordance with the manufacturer’s protocols.

***Genetic risk profiling***

In this study, we leveraged whole-genome sequencing data from the UK Biobank encompassing all 500,000 participants to assess the effect of genetic susceptibility^2^. The polygenic risk score (PRS) for IBD and CD was generated using the SCT approach as implemented in the R package “bigsnpr”. SNP effect sizes were obtained from a meta-analyzed genome-wide association study (GWAS) summary statistics provided by the VA Million Veteran Program^3^. A comprehensive grid search was conducted across approximately 1,400 parameter settings, covering a broad spectrum of linkage disequilibrium (LD) clumping criteria (r² thresholds ranging from 0.01 to 0.95 with clumping windows of 250-500 kb) and GWAS *P*-value cutoffs (from 1×10^-8^ to 0.1)^4^. To combine candidate scores, elastic net regression with 10-fold cross-validation was applied, and the optimal model was selected by jointly considering model simplicity and predictive accuracy^4^. Participants were categorized into high or low genetic risk groups for IBD and CD based on the median value of PRS. In the joint analysis, participants were classified into four groups according to the median of UPF intake and PRS for CD, with the low UPF intake and low PRS group serving as the reference.

***Identification of metabolite modules through WGCNA***

The WGCNA is a method used for computing co-expression networks, which have recently been used to integrate omics data18. By setting soft-threshold power (ranging from 1-20) to limit the range of correlation coefficients between features, we applied WGCNA to metabolites filtered by the elastic net model. The identified metabolites were clustered into modules by hierarchical clustering using Euclidean distance and labelled with distinct colors. After metabolite clustering, we calculated the combined z-score of metabolites for each identified module, and further assessed the association between the module’s combined z-score and incident IBD and CD using Cox regression models. To ensure the consistency and stability of the clustered metabolites, we performed WGCNA in the discovery cohort and applied the clustered metabolite modules for subsequent analyses. In the current analysis, the scale-free topology model fit (R^2^) reached the highest as 0.76 for metabolomics analysis when the soft thresholding power was 6. We used the minimum module size of 3 for metabolites. Based on the cluster-level results, we chose the module that showed the strongest association with incident CD.

***Selection for instrument variables and global GWAS for a single metabolite***

We leveraged the summary-level data from the global Genome-Wide Association Study (GWAS) analysis of 233 circulating metabolites among 136,016 participants from 33 cohorts based on the same NMR metabolomics platform from Nightingale Health, which provides all-encompassing genome-wide characterization of circulating metabolic biomarkers^5^. The metabolic trait distributions were adjusted for age, sex, principal components, and relevant study-specific covariates. We identified the single nucleotide polymorphism (SNP) associated with target metabolite (docosahexaenoic acid [DHA] in current analysis) to select independent instrument variables (IVs) for target metabolite based on the following criteria: (1) SNPs should be associated with circulating DHA at a genome-wide significance level with a threshold of *P* < 5 × 10^−8^; (2) SNPs should not in linkage disequilibrium (LD) with other genetic instruments for the same exposure (defining as r^2^ < 0.01).

Summary-level GWAS data for CD and UC were available in the UK Biobank, the FinnGen study (10^th^ release), and the International Inflammatory Bowel Disease Genetics Consortium (IIBDGC). We used METAL as a tool for the meta-analysis of the three GWAS. In this study, summary statistics of genetic associations in UKB were extracted from GWAS conducted by the Lee lab. IBD cases (including CD and UC) were identified through the inpatient data. The estimates of gene-disease association were obtained by logistic regression with adjustment for the genetic principal components, sex, and birth year. The FinnGen study is a large nationwide cohort study launched in 2017, which combined genetic data from Finnish biobanks and digital health record data from Finnish health registries^6^. IBD cases were identified based on the registry in the social insurance institution, with diagnostic information based on ICD codes. Genome-wide association analyses for each trait were adjusted for sex, age, genetic components, and genotyping batch. IIBDGC brings together genome-wide genotyping data and whole-genome sequencing data for over 75,000 patients with IBD^7^. Diagnosis of IBD in IIBDGC was based on accepted radiologic, endoscopic, and histopathologic evaluations. The genetic associations were obtained from logistic regression adjusted for age, sex, and genetic principal components. We employed European ancestry summary-level statistics, including data for CD (5,956 cases and 14,927 controls) and UC (6,968 cases and 20,464 controls).

***Mendelian randomization, colocalization, and G-E interaction analysis***

To assess the consistency of the results and account for horizontal pleiotropy, we performed sensitivity analyses for MR analysis using methods including the weighted median, MR-Egger, MR pleiotropy residual sum and outlier (MR-PRESSO), and leave-one-out MR analysis. The weighted median method provides consistent estimates when ≥50% of the instrumental variables are valid. This method works by taking the median of the estimates from all the instruments, ensuring that if the majority of instruments are valid, the estimate will still be accurate even if some instruments are invalid or biased^8^. MR-Egger regression helps adjust for horizontal pleiotropic effects, which occur when genetic variants influence the outcome through pathways other than the exposure of interest^9^. This regression method offers unbiased causal estimates, although the estimates may be less precise and may have lower statistical power, especially when the number of instruments is small, or the pleiotropic effects are weak^9^. MR-PRESSO, a method designed to identify and correct for horizontal pleiotropic outliers, can identify and correct for horizontal pleiotropic outliers^10^.

Furthermore, we performed colocalization analysis to identify potentially shared causal variants between key metabolites and incident IBD, with the R package “coloc”^11^. Five different posterior probabilities are reported in colocalization analysis, corresponding to five hypotheses: (a) no shared causal variants for either of the two traits (H0); (b) a causal variant for gene expression only (H1); (c) a causal variant for disease risk only (H2); (d) distinct causal variants for both traits (H3); and (e) a same shared causal variant for both traits (H4). For colocalization of GWAS data, the colocalization region windows were ±1000kb according to a previous publication^12^. The prior probabilities that the causal variants are associated with only trait 1 (key metabolite), only trait 2 (IBD), and both are set at 10^-4^, 10^-4,^ and 10^-5,^ respectively. Posterior probabilities for hypothesis H4 (shared causal variant) were calculated, with a posterior probability (PPH4) greater than 0.70 considered strong evidence for colocalization, suggesting a shared genetic influence on both traits, with its cutoff corresponding to a false discovery rate (FDR) of <5%. HyPrColoc is a Bayesian algorithm designed for multi-trait statistical colocalization using GWAS summary statistics, enabling large-scale trait analysis. It can efficiently assess if multiple traits share a causal variant via the Posterior Probability of Full Colocalization (PPFC), with linear computational cost allowing rapid analysis of hundreds of traits (e.g., ~1 second for 100 traits in a 1000-SNP region). When PPFC is low (indicating non-universal colocalization), it uses a branch-and-bound clustering algorithm to group traits into subsets, each satisfying colocalization criteria to identify distinct shared causal variants. It also features a parsimonious prior setup (two interpretable parameters) that supports sensitivity analyses, making it practical for real-world studies, such as integrating disease and molecular traits to prioritize candidate causal genes and risk regions. Pair-Wise Conditional analysis and Colocalisation analysis (PWCoCo) were conducted to scan non-primary signals in the region which colocalize^13^. To investigate the gene-environment (G-E) interaction between genotype of key genetic variants (G) and UPF intake (E), we constructed a regression model including genotype, UPF intake, their interaction term (genotype × UPF intake), and relevant covariates in the fully-adjusted model^14^. The interaction effect was assessed by testing the statistical significance of the interaction term’s regression coefficient (two-tailed *P*-value < 0.05 was considered significant).

### **Supplementary Figures**

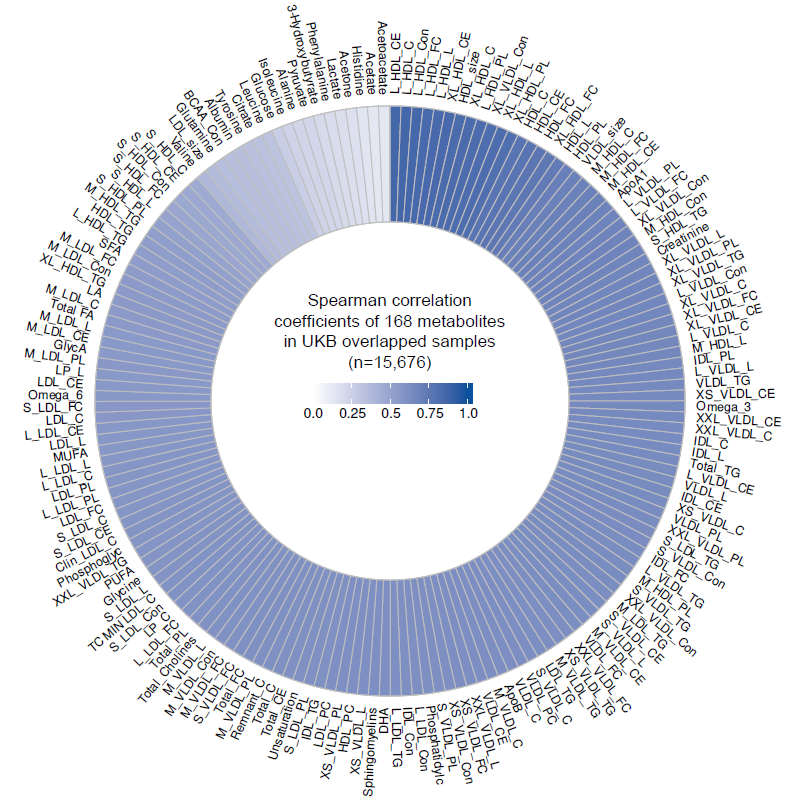

**Figure S1.** Spearman correlation coefficients of 168 metabolites in UKB baseline and follow-up measurements in the overlapped samples.

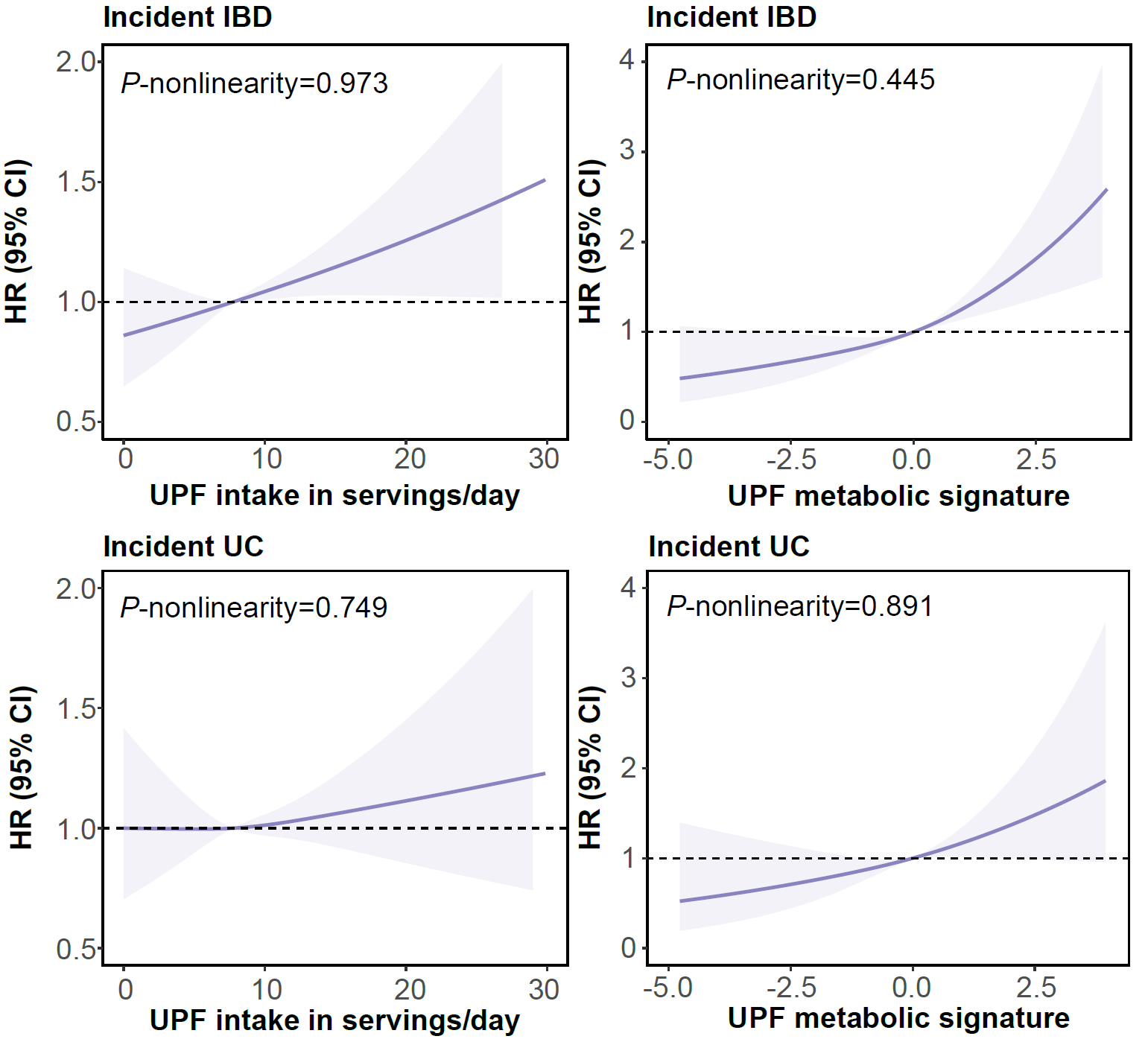

**Figure S2.** The dose-response association in the survival analysis between UPF intake, UPF metabolic score, and incident IBD and UC. IBD, inflammatory bowel disease; UC, ulcerative colitis; UPF, ultra-processed food.

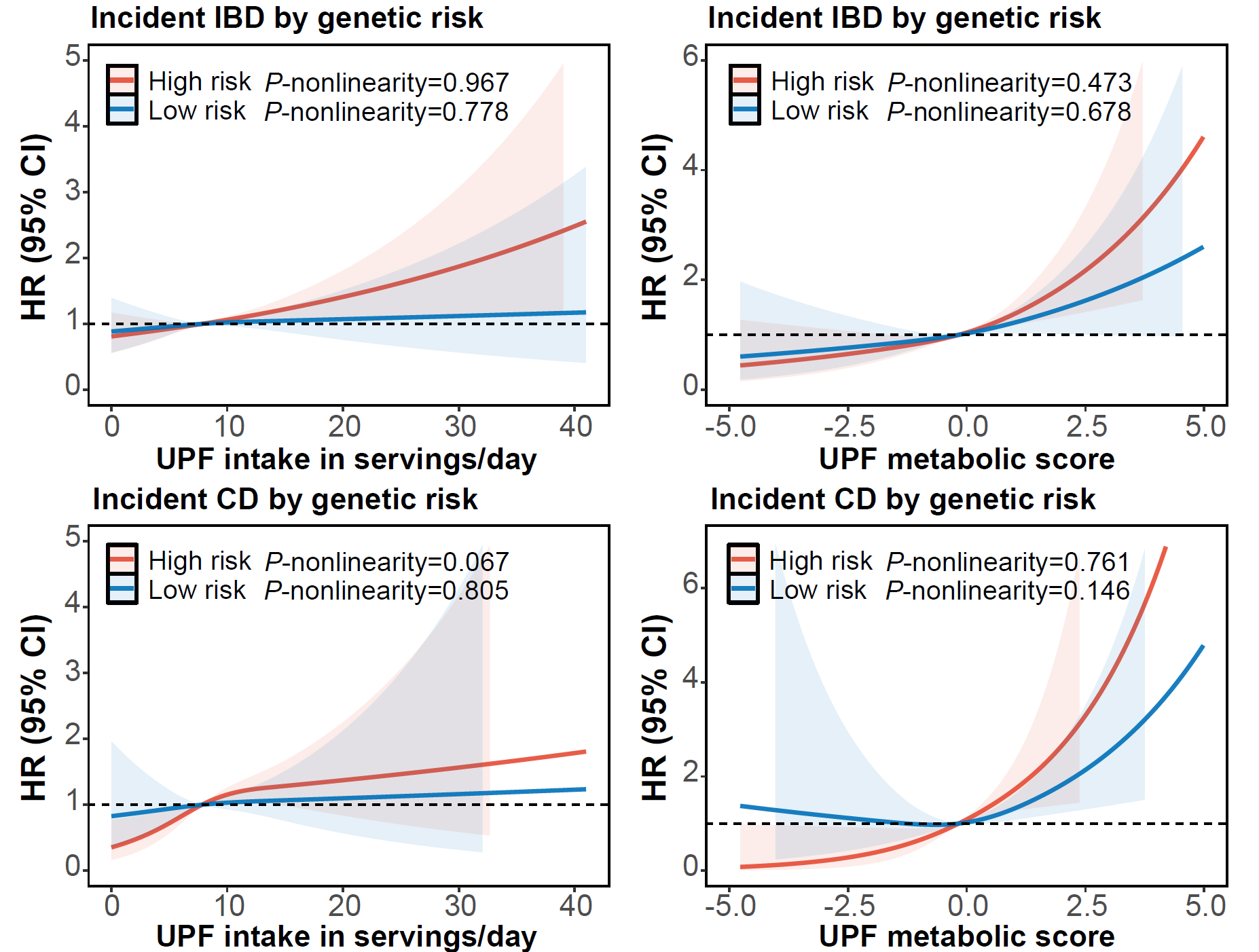

**Figure S3.** The dose-response association between UPF intake, UPF metabolic score, and incident IBD and CD, stratified by genetic risk. IBD, inflammatory bowel disease; CD, Crohn’s disease; UPF, ultra-processed food.

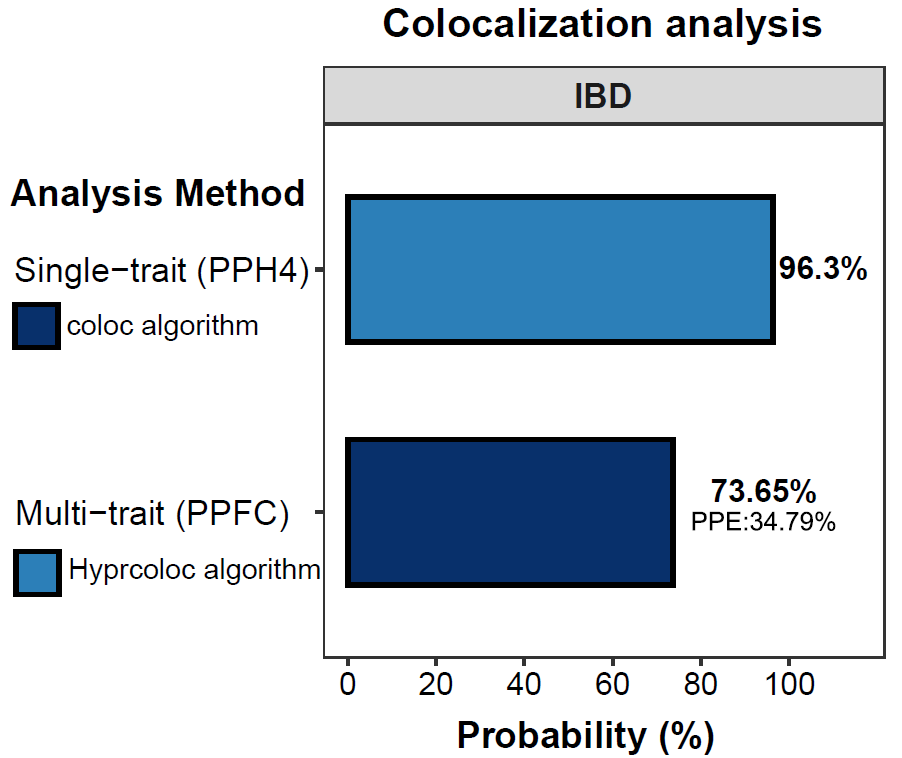

### **Figure S4.** Colocalization analysis of shared genetic variants of DHA and IBD.

Results of the colocalization analysis applying two methods, indicating evidence for shared causal variation between circulating DHA levels and incident IBD. DHA, docosahexaenoic acid; IBD, inflammatory bowel disease; PPFC, posterior probability of full colocalization; PPE, proportion of PPFC explained by the listed SNP; PPH4, posterior probability of hypothesis 4.

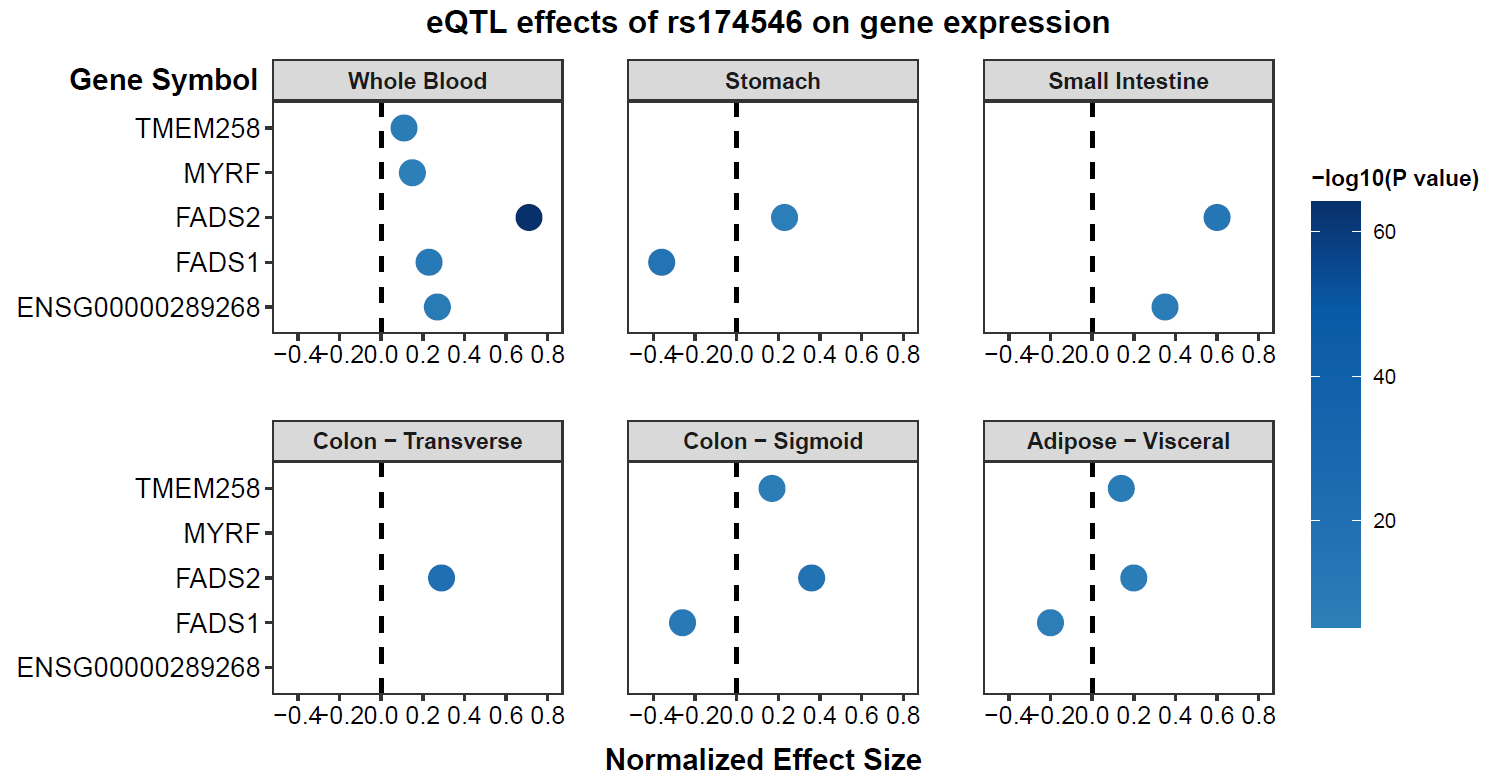

### **Figure S5.** The expression quantitative trait locus (eQTL) effects of rs174546 on the expression of genes (*TMEM258*, *MYRF*, *FADS2*, *FADS1*, and ENSG00000289268) across various tissues based on GTEx v8. The horizontal axis denotes the Normalized Effect Size. The color bar on the right represents −log_10_(*P*-value), with darker colors indicating smaller *P* values and thus higher statistical significance of the eQTL associations.

### **Supplementary Tables**

### **Table S1.** Items for ultra-processed food in the UK Biobank, Whitehall II, and ONE-IBD

| **Code** | **Food item** | **Portion size**  **g/serving ^a^** | **Energy**  **KJ/100 g ^b^** |
| --- | --- | --- | --- |
| **UK Biobank** | | | |
| 100160 | Low-calorie drink intake | 330 | 43 |
| 100170 | Fizzy drink intake: carbonated (fizzy) drinks | 330 | 174 |
| 100180 | Squash intake | 250 | 159 |
| 100250 | Instant coffee intake | 190 | 8 |
| 100380 | Intake of artificial sweetener added to coffee ^c^ | 6 | 200 |
| 100500 | Intake of artificial sweetener added to tea ^d^ | 6 | 200 |
| 100530 | Flavoured milk intake | 250 | 270 |
| 100720 | Fortified wine intake | 50 | 481 |
| 100730 | Spirits intake | 23 | 919 |
| 100770 | Porridge intake | 203.5 | 195 |
| 100800 | muesli intake | 100 | 1540 |
| 100810 | Oat crunch intake | 100 | 1639 |
| 100820 | Sweetened cereal intake | 38 | 1632 |
| 100830 | Plain cereal intake | 30 | 1601 |
| 100840 | Bran cereal intake | 50 | 1406 |
| 100850 | Whole-wheat cereal intake | 44 | 1474 |
| 100860 | Other cereal intake | 44 | 1566 |
| 101090 | Bap intake | 90 | 1065 |
| 101160 | Bread roll intake | 60 | 1084 |
| 101230 | Naan bread intake | 160 | 1206 |
| 101250 | Crispbread intake | 10 | 1591 |
| 101260 | Oatcakes intake | 13 | 1737 |
| 101270 | Other bread intake | 45 | 1661 |
| 101310 | Number of bread slices with butter/margarine | Thin 10 medium 12 thick 15 | 3061 |
| 101350 | Number of baguettes with butter/margarine |  |  |
| 101390 | Number of baps with butter/margarine |  |  |
| 101430 | Number of bread rolls with butter/margarine |  |  |
| 101470 | Number of crackers/crispbreads with butter/margarine |  |  |
| 101510 | Number of oatcakes with butter/margarine |  |  |
| 101550 | Number of other bread types with butter/margarine |  |  |
| 101970 | Double-crusted pastry intake | 60 | 1060 |
| 101980 | Single-crust pastry intake | 30 | 1310 |
| 101990 | Crumble intake | 70 | 924 |
| 102000 | Pizza intake | 150 | 1036 |
| 102010 | Pancake intake | 110 | 1065 |
| 102020 | Scotch pancake intake | 41 | 1138 |
| 102030 | Yorkshire pudding intake | 25 | 881 |
| 102040 | Indian snacks intake | 40 | 975 |
| 102050 | Croissant intake | 60 | 1563 |
| 102060 | Danish pastry intake | 110 | 1441 |
| 102070 | Scone intake | 48 | 1378 |
| 102120 | Ice-cream intake | 120 | 1229 |
| 102140 | Milk-based pudding intake | 200 | 494 |
| 102150 | Other milk-based pudding intake | 60 | 518 |
| 102170 | Soya dessert intake | 125 | 309 |
| 102180 | Fruitcake intake | 70 | 1478 |
| 102190 | Cake intake | 60 | 1617 |
| 102200 | Doughnut intake | 60 | 1414 |
| 102210 | Sponge pudding intake | 120 | 1116 |
| 102220 | Cheesecake intake | 110 | 1231 |
| 102230 | Other dessert intake | 60 | 696 |
| 102260 | Chocolate bar intake | 50 | 2081 |
| 102270 | White chocolate intake | 50 | 2212 |
| 102280 | Milk chocolate intake | 50 | 2177 |
| 102290 | Dark chocolate intake | 50 | 2273 |
| 102300 | Chocolate-covered raisin intake | 25 | 1159 |
| 102310 | Chocolate sweet intake | 36 | 1922 |
| 102320 | Diet sweet intake | 18 | 975 |
| 102330 | Sweets intake | 36 | 1793 |
| 102340 | Chocolate-covered biscuits intake | 17 | 2076 |
| 102350 | Chocolate biscuits intake | 24 | 2071 |
| 102360 | Sweet biscuits intake | 17 | 1842 |
| 102370 | Cereal bar intake | 28 | 1525 |
| 102380 | Other sweets intake sweet snacks | 40 | 1793 |
| 102460 | Crisp intake | 40 | 2186 |
| 102470 | Savoury biscuits intake | 40 | 2168 |
| 102480 | Cheesy biscuits intake | 40 | 1366 |
| 102500 | Other savoury snack intake | 40 | 2099 |
| 102530 | Powdered/instant soup intake | 200 | 270 |
| 102760 | Snackpot intake (snack pot, noodles/rice | 280 | 1541 |
| 102770 | Couscous intake | 150 | 1440 |
| 102850 | Low-fat cheese spread intake | 40 | 733 |
| 102860 | Cheese spread intake | 15 | 1106 |
| 103010 | Sausage intake | 30 | 1218 |
| 103050 | Crumbed or deep-fried poultry intake (chicken or turkey in breadcrumbs or deep-fried | 100 | 1111 |
| 103070 | Bacon intake | 46 | 891 |
| 103080 | Ham intake | 23 | 891 |
| 103260 | Vegetarian sausages/burgers intake | 90 | 748 |
| 103280 | Quorn intake | 90 | 389 |
| 103290 | Other vegetarian alternative intake | 90 | 1386 |
| 104000 | Baked bean intake | 135 | 335 |
| 104020 | Fried potatoes intake | 180 | 796 |
| 104050 | Mashed potato intake | 60 | 438 |
| **Whitehall II Study** | |  |  |
| XBROBRD | Bread: Avg brown bread/rolls | 60 | 1084 |
| XCRISBRD | Bread: Avg crispbread | 10 | 1591 |
| XWHIBRD | Bread: Avg white bread/rolls | 60 | 1084 |
| XWHOLBRD | Bread: Avg whole meal bread/rolls | 60 | 1084 |
| XBRAN | Cereals: Avg All-Bran, Bran Flakes | 50 | 1406 |
| XCFLAK | Cereals: Avg Corn Flakes, Rice Krispies | 30 | 1601 |
| XFROS | Cereals: Avg Frosties, Ricicles, etc | 30 | 1601 |
| XMUES | Cereals: Avg Muesli, Fruit n Fibre | 100 | 1540 |
| XSHRED | Cereals: Avg Shredded w, Weetabix | 44 | 1474 |
| XPORRI | Cereals: Avg porrige, Readybrek | 203.5 | 195 |
| XDRMILK | Dairy: Avg dried milk | 3 | 1482 |
| XHORLI | Drink: Avg Horlicks, Ovaltine | 20 | 1553 |
| XCOFFWH | Drink: Avg coffee whitener | 3 | 2254 |
| XFIZZY | Drink: Avg fizzy soft drinks | 330 | 174 |
| XSQUASH | Drink: Avg fruit squash or cordial | 250 | 159 |
| XLOWCAL | Drink: Avg low cal, diet fiz drinks | 330 | 43 |
| XPORT | Drink: Avg port, cherry, vermouth | 50 | 481 |
| XFJUICE | Drink: Avg real fruit juice | 250 | 164 |
| XSPIRITS | Drink: Avg spirits | 23 | 919 |
| XHARDMAR | Fat: Avg hard margarine | 12 | 3601 |
| XLFSPREA | Fat: Avg low fat spread | 12 | 3601 |
| XSOFTMAR | Fat: Avg other soft margarine | 12 | 3601 |
| XPOLYUNS | Fat: Avg polyunsat margarine | 12 | 3601 |
| XFISHFIN | Fish: Avg fish fingers, fish cakes | 28 | 994 |
| XBATFIS | Fish: Avg fried fish in batter | 100 | 1031 |
| XBACON | Meat: Avg bacon | 46 | 891 |
| XBEEFBU | Meat: Avg beef burgers | 105 | 1206 |
| XCORNBF | Meat: Avg corned beef, luncheon. | 120 | 860 |
| XHAM | Meat: Avg ham | 23 | 891 |
| XLIVER | Meat: Avg liver, liver pate | 70 | 734 |
| XSAUSAG | Meat: Avg sausages | 30 | 1218 |
| XSAVPIE | Meat: Avg savory pies | 140 | 1514 |
| XLASAGNE | Pasta: Avg lasagne | 420 | 800 |
| XPIZZA | Pasta: Avg pizza | 150 | 1036 |
| XQUICHE | Pasta: Avg quiche | 140 | 1488 |
| XCHIPS | Potato: Avg chips or French fries | 180 | 1174 |
| XROASPOT | Potato: Avg roast potatoes | 180 | 796 |
| XMARMITE | Sauces: Avg Marmite, Bovril | 62 | 763 |
| XVINAIGR | Sauces: Avg French dressing, vinaigre | 15 | 1902 |
| XPICKLES | Sauces: Avg pickles, chutney | 40 | 1202 |
| XMAYO | Sauces: Avg salad cream | 20 | 1440 |
| XSAUCE | Sauces: Avg sauces - white, gravy | 62 | 626 |
| XKETCHU | Sauces: Avg tomato ketchup | 20 | 489 |
| XBISCUIT | Sweets/snacks: Avg biscuits | 17 | 1842 |
| XBUNS | Sweets/snacks: Avg buns and pastries | 110 | 1441 |
| XCAKES | Sweets/snacks: Avg cakes | 60 | 1617 |
| XCHOC | Sweets/snacks: Avg chocolates, choc bars | 50 | 2081 |
| XCOCOA | Sweets/snacks: Avg cocoa, hot chocolate | 6 | 1301 |
| XCRACKER | Sweets/snacks: Avg cream crack, cheese bisc | 7 | 1746 |
| XCRISPS | Sweets/snacks: Avg crisps, packet snacks | 40 | 2186 |
| XTARTS | Sweets/snacks: Avg fruit pies, tarts, crumble | 110 | 822 |
| XICECREA | Sweets/snacks: Avg ice cream, choc ices | 120 | 1229 |
| XJAM | Sweets/snacks: Avg jam, marmalade, honey | 15 | 1114 |
| XMILKPUD | Sweets/snacks: Avg milk puddings | 200 | 494 |
| XSPONGE | Sweets/snacks: Avg sponge puddings | 120 | 1116 |
| XSWEETS | Sweets/snacks: Avg sweets, toffees, mints | 40 | 2099 |
| XTVP | Veg: Avg soya meat, TVP, vegebur | 90 | 807 |
| XSOYMLK | Veg: Avg soya milk | 585 | 182 |
| XTOFU | Veg: Avg tofu or soya bean curd | 90 | 304 |
| **ONE-IBD Study** | | | |
| 1 | Sweets | 3-10.5 | 429 |
| 2 | Ice cream | 36 | 221 |
| 3 | Chips | 8-60 | 370 |
| 4 | Pizza | 61 | 266 |
| 5 | Sugar-sweetened beverages | - | 42 |
| 6 | Artificial-sweetened beverages | - | 2 |
| 7 | Salt | 6 | 0 |
| 8 | Processed oil from plants | 14.5 | 899 |
| 9 | Processed oil from animals | 14.5 | 227 |

^a^ Portion size used here was according to UK McCance and Widdowson’s *The Composition of Foods 6th edition (2002) and its supplements* ^1,2^. In ONE-IBD, some food items have multiple portion sizes due to different categories of food.

^b^ Energy for each food used here was according to *UK McCance and Widdowson’s The Composition of Foods 6th edition (2002) and its supplements.*^3^

^c^ Coffee here included Instant coffee (100250), Filtered coffee (100270), Cappuccino (100290), Latte (100300), Espresso (100310), Other coffee type (100330).

^d^ Tea here included Standard tea (100400), Rooibos tea (100410), Green tea (100420), Herbal tea (100430), Other tea (100440).

### **Table S2.** Diagnostic codes of inflammatory bowel disease

| **Sources** | **Diagnostic codes** | **Code set** |
| --- | --- | --- |
| Primary care data | Read code (v2) | **CD**: J40.., J400., J4000, J4001, J4002, J4003, J4004, J4005, J400z, J401., J4010, J4011, J4012, J401z, J08z9, J402., Jyu40, J40z., N0311;  **UC**: J4100, J411., J412., J413., J4103, J4102, J41y0, J438., J410., J41y., J41yz, J41z., Jyu41, J41.., J4101, J4104, J410z, N0310. |
|  | Read code (v3) | **CD**: J5109, XE0ae, XE2QL, Xa0lh, Xa8Eh, J400., J4000, J4001, J4002, J4003, J4004, J400z, X302r, X302s, X302t, X302u, XaK6C, J401., J4010, J4011, J401z, X3050, XE0af, XaK6D, J08z9, J400., J4000, J4002, J4010, J4011, J402., Jyu40, X20Pq, X300J, X301b, X302r, X302t, J40z., J5109, XE2QL, Xa0lh, N0311, N0453, X701t, X7021, X702C;  **UC**: J410., X302z, X304H, XE0ae, XE0ag, Xa8Eh, XaK6E, J4100, X302s, X3030, XaB5z, XaYzX, J4103, X303e, J41y0, XaZ2j, J41y., J41yz, J41z., Jyu41, X302y, J41.., J410., J410z, X302y, XE0ag, XaK6E, N0310, N0454, X7021, X702C. |
| Hospital inpatient data | ICD-9  ICD-10 | **CD**: 555; **UC**: 556.  **CD**: K50; **UC**: K51. |
| Death registry data | ICD-10 | **CD**: K50; **UC**: K51. |

ICD, International Classification of Diseases; CD, Crohn's disease; UC, ulcerative colitis.

### **Table S3.** Definition of covariates

| **Variable** | **Description** |
| --- | --- |
| **UK Biobank** | |
| Age at recruitment | This is a derived variable based on date of birth and date of attending an initial assessment center and refers to the age of the participant on the day they attended an Initial Assessment Centre, truncated to whole year.  (response: as continues variable) |
| Sex | A mixture of the sex the National Health Service had recorded for the participant and self-reported sex (response: categorical variable “**Female**”, “**Male**”) |
| Ethnicity | Self-reported: “What is your ethnic background”. We classified the responses into: **White** (White) and **Others** (Mixed, Asian or Asian British, Black or Black British, Chinese, and other ethnic group) |
| Education | Self-reported: “Which of the following qualifications do you have” We classified the responses into: **College** (College or University degree) and **Below college** (A levels/AS levels or equivalent, O levels/GCSEs or equivalent, CSEs or equivalent, NVQ or HND or HNC or equivalent, other professional qualifications, e.g.: nursing, teaching, and none of the above) |
| Townsend deprivation index (TDI) | The higher, the more socioeconomic deprivation one was suffering. TDI was derived according to the unemployment rate, the percentage of overcrowded households, the percentage of people without cars, and the percentage of people without houses for each area in the UK, and baseline TDI calculated immediately before participant joining UK Biobank based on the preceding national census output areas. Each participant was assigned a score corresponding to the output area in which their postcode is located ^a^.  (response: as continues variable) |
| Smoking status | Self-reported current/past smoking status of the participant. We classified the responses into: **Never smoked** (Never) and **Former or current smoker** (Previous or current) |
| Physical activity | Self-reported: UK Biobank physical activity questionnaire. We classified the responses into **Adequate** (150 minutes moderate activity per week OR ≥ 75 minutes vigorous activity per week OR equivalent combination OR moderate physical activity at least 5 days a week or vigorous activity once a week) and **Inadequate** (below adequate level) recommended by the American Heart Association. |
| Body mass index (BMI) | BMI value is constructed from height and weight measured during the initial assessment center visit. Relevant variable was measured by trained staff.  (response: as continuous variable) |
| Total energy intake | Total energy from overall diet estimating from the mean intake of the 24-h WebQ |
| Total sugar intake | Sugar estimating from the mean intake of the 24-h WebQ |
| **Whitehall II Study** | |
| Year of birth | Birth year of participants (response: as continues variable) |
| Sex | Self-reported sex (response: as categorical variable “**Female**”, “**Male**”) |
| Ethnicity | Self-reported: “What is your ethnic background”. The answer is **White** and **Non-white** |
| Townsend deprivation index (TDI) | Same as the UKB definition |
| Smoking status | Self-reported current/past smoking status of the participant. We classified the responses into: **Never smoked**, **Previous smoker**, and **Current smoker**. |
| Total energy intake | Estimating from the food frequency questionnaire |
| Body mass index (BMI) | BMI value is constructed from height and weight measured at initial clinical assessment  (response: as continuous variable) |
| **ONE-IBD Study** |  |
| Age at recruitment | This is a derived variable based on date of birth and date of attending an initial assessment center and refers to the age of the participant on the day they attended an Initial Assessment Centre, truncated to whole year.  (response: as continues variable) |
| Sex | The mix of biological and self-reported sex (response: categorical variable “**Female**”, “**Male**”) |
| Ethnicity | Self-reported: “What is your ethnic background”. We classified the responses into: **Han** and **Others** (other ethnic groups in East Asia) |
| Education | Self-reported: “Which of the following qualifications do you have” We classified the responses into: **College** (postgraduate, college or university degree) and **Below college** (high school, middle school, primary school, or none of the above) |
| Body mass index (BMI) | BMI value is constructed from height and weight measured during the initial assessment center visit. Relevant variable was measured by trained staff.  (response: as continuous variable) |
| Smoking status | Self-reported current/past smoking status of the participant. We classified the responses into: **Never smoked** (Never) and **Former or current smoker** (Previous or current) |
| Physical activity | Self-reported physical activity questionnaire, and calculated by MET scores. We classified the responses into Adequate (150 minutes moderate activity per week) and Inadequate (below adequate level) recommended by the American Heart Association. |
| Total energy | Total energy from overall diet estimating from the mean intake of the Food Frequency Questionnaire (FFQ) |

^a^ Reference: Blane D, Townsend P, Phillimore P, et al. Health and Deprivation: Inequality and the North. British Journal of Sociology 1987;40:344.

### **Table S4.** Missing rate of covariates in UK Biobank, Whitehall II study, and ONE-IBD

| **Covariates** ^a^ | **UK Biobank**  **(discovery cohort)** | | **UK Biobank**  **(internal validation)** | | **Whitehall II**  **(external validation)** | | **ONE-IBD**  **(external validation)** | |
| --- | --- | --- | --- | --- | --- | --- | --- | --- |
|  | **Number** | **Missing Rate (%)** | **Number** | **Missing Rate (%)** | **Number** | **Missing Rate (%)** | **Number** | **Missing Rate (%)** |
| Age at recruitment | 0 | 0 | 0 | 0 | 0 | 0 | 0 | 0 |
| Sex | 0 | 0 | 0 | 0 | 0 | 0 | 0 | 0 |
| Ethnicity | 0 | 0 | 0 | 0 | 28 | 0.35 | 0 | 0 |
| Townsend deprivation index (TDI) | <20 | <0.01 | 118 | 0.13 | 385 | 4.88 | - | - |
| Smoking status | <20 | <0.01 | 224 | 0.25 | 563 | 7.13 | 106 | 14.1 |
| Physical activity | 0 | 0 | 0 | 0 | - | - | 26 | 3.46 |
| Body mass index (BMI) | 22 | 0.22 | 214 | 0.23 | 259 | 3.28 | 0 | 0 |
| Total energy intake | 0 | 0 | 0 | 0 | 0 | 0 | 0 | 0 |
| Total sugar intake | 0 | 0 | 0 | 0 | 0 | 0 | 0 | 0 |

^a^ Continuous variables were imputed with the median. Categorical variables were filled in with the most frequent category (missing rate <5%) or using a missing indicator (missing rate≥5%).

### **Table S5.** Genetic instrument variables selected for Mendelian randomization analysis.

| **Exposure** | **Outcome** | **SNP** | **Chr** | **Position ^a^** | **Effect allele** | **Other allele** |
| --- | --- | --- | --- | --- | --- | --- |
| DHA | CD | rs78905952 | 1 | 176784526 | T | G |
| DHA | CD | rs4846923 | 1 | 230307222 | T | G |
| DHA | CD | rs4601123 | 6 | 11050803 | C | G |
| DHA | CD | rs10822160 | 10 | 65112796 | T | G |
| DHA | CD | rs10885997 | 10 | 118397971 | A | G |
| DHA | CD | rs489172 | 11 | 60637164 | T | C |
| DHA | CD | rs139907587 | 11 | 60786289 | T | C |
| DHA | CD | rs509936 | 11 | 60858031 | A | G |
| DHA | CD | rs3019200 | 11 | 61249383 | A | C |
| DHA | CD | rs3741259 | 11 | 61282350 | T | C |
| DHA | CD | rs117566820 | 11 | 61285641 | A | G |
| DHA | CD | rs75444393 | 11 | 61359218 | T | C |
| DHA | CD | rs139343543 | 11 | 61394647 | T | C |
| DHA | CD | rs60399071 | 11 | 61443155 | A | G |
| DHA | CD | rs533665305 | 11 | 61463351 | T | C |
| DHA | CD | rs2299652 | 11 | 61525817 | T | C |
| DHA | CD | rs149570869 | 11 | 61526688 | A | G |
| DHA | CD | rs12418216 | 11 | 61549399 | A | T |
| DHA | CD | rs76368178 | 11 | 61568827 | T | C |
| DHA | CD | rs174546 | 11 | 61569830 | T | C |
| DHA | CD | rs72643557 | 11 | 61579427 | T | C |
| DHA | CD | rs72920193 | 11 | 61602572 | T | C |
| DHA | CD | rs11605884 | 11 | 61630133 | T | C |
| DHA | CD | rs111509315 | 11 | 61728651 | A | G |
| DHA | CD | rs143385562 | 11 | 61788314 | T | C |
| DHA | CD | rs184129994 | 11 | 61818622 | T | C |
| DHA | CD | rs12226389 | 11 | 61823630 | T | C |
| DHA | CD | rs2229738 | 11 | 68562328 | T | C |
| DHA | CD | rs600626 | 11 | 75455309 | A | G |
| DHA | CD | rs10468017 | 15 | 58678512 | T | C |
| DHA | CD | rs261334 | 15 | 58726744 | C | G |
| DHA | CD | rs12928099 | 16 | 15150505 | A | C |
| DHA | CD | rs56228609 | 16 | 56987765 | T | C |
| DHA | CD | rs4646368 | 17 | 17465458 | A | G |
| DHA | CD | rs8107974 | 19 | 19388500 | A | T |
| DHA | CD | rs182611493 | 19 | 19458388 | A | G |
| DHA | CD | rs35336243 | 19 | 45442519 | A | T |
| DHA | CD | rs838147 | 19 | 49246866 | A | G |
| DHA | CD | rs4806498 | 19 | 54674742 | T | C |

^a^ The position on the chromosome identified by data from Gtexv8.

### **Table S6.** Baseline characteristics of participants with available dietary and metabolomic data in three cohorts.

| **Characteristics** | **UK Biobank** | | **Whitehall II** | **ONE-IBD Study** | |
| --- | --- | --- | --- | --- | --- |
|  | **Discovery cohort** | **Internal validation** | **External validation** | **Inflammatory bowel disease** | **Healthy controls** |
|  | n=10,229 | n=91,306 | n=7,893 | n=392 | n=360 |
| Age (mean (SD)) | 57.05 (7.33) | 56.20 (7.95) | 53.32 (6.07) | 34.46 (13.39) | 34.52 (10.07) |
| Sex (%) |  |  |  |  |  |
| Female | 5,200 (50.8) | 50,060 (54.8) | 2,411 (30.5) | 103 (26.3) | 220 (61.1) |
| Male | 5,029 (49.2) | 41,246 (45.2) | 5,482 (69.5) | 289 (73.7) | 140 (38.9) |
| Ethnicity (%) ^a^ |  |  |  |  |  |
| The majority | 10,069 (98.4) | 87,805 (96.2) | 7,211 (91.4) | 386 (98.5) | 349 (96.9) |
| Others | 160 (1.6) | 3,501 (3.8) | 682 (8.6) | 6 (1.5) | 9 (2.5) |
| TDI (mean (SD)) | -2.18 (2.60) | -1.62 (2.86) | -0.07 (0.95) | - | - |
| Education (%) |  |  |  |  |  |
| College and above | 4,981 (48.7) | 37,704 (41.3) | - | 222 (56.6) | 336 (93.3) |
| Below college | 5,248 (51.3) | 53,602 (58.7) | - | 170 (43.4) | 24 (6.7) |
| BMI (mean (SD)) | 26.67 (4.41) | 26.97 (4.64) | 25.61 (4.04) | 20.86 (4.34) | 22.79 (4.28) |
| Smoking (%) |  |  |  |  |  |
| Never | 6,120 (59.8) | 51,905 (56.8) | 3,414 (43.3) | 172 (43.9) | 310 (86.1) |
| Previous or current | 4,109 (40.2) | 39,401 (43.2) | 3,916 (49.6) | 114 (29.1) | 50 (13.9) |
| Unknown |  |  |  | 106 (27.0) | 0 (0.0) |
| Physical activity (%) |  |  |  |  |  |
| Adequate ^b^ | 7,329 (71.6) | 65,349 (71.6) | - | 204 (52.8) | 202 (56.2) |
| Inadequate | 2,900 (28.4) | 25,957 (28.4) | - | 162 (41.3) | 158 (43.8) |
| Dietary sugar intake, grams per day (mean (SD)) | 127.08 (44.46) | 124.51 (47.91) | - | 310.30 (119.50) | 330.17 (140.78) |
| Energy intake, kJ/day (mean (SD)) | 8,620.03 (2123.52) | 8,453.12 (2277.49) | 10,258.18 (2731.69) | 9358.88 (3219.28) | 9028.60 (3139.08) |
| UPF serving per day (mean (SD)) | 8.63 (4.19) | 8.55 (4.51) | 10.55 (4.32) | - | - |
| UPF grams per day (mean (SD)) | 672.60 (409.85) | 670.53 (435.52) | 701.85 (314.18) | 134.16 (207.33) | 140.08 (145.63) |
| UPF energy per day (mean (SD)) | 3,686.73 (1693.14) | 3,619.39 (1782.00) | 5,875.32 (2296.40) | 319.84 (167.44) | 314.55 (121.66) |
| UPF energy proportion per day (mean (SD)) | 0.41 (0.14) | 0.41 (0.15) | 0.56 (0.12) | 0.14 (0.08) | 0.15 (0.07) |

BMI, body mass index; TDI, Townsend deprivation index; SD, standard deviation; UPF, ultra-processed food.

^a^ Ethnicity was categorized according to cohort-specific population structure. In the UK Biobank and Whitehall II Study, the majority refers to the White/European ancestry, while in the ONE-IBD Study, the majority refers to the Han Chinese group.

^b^ Adequate physical activity level meets at least one of the following criteria: a. doing moderate or vigorous-intensity activity at least 150 minutes in metabolic equivalent minutes per week (MET-minutes/week); or b. more than 5 days/week with over 10 minutes moderate physical activity; c. more than one day/week with over 10 minutes vigorous physical activity. Other circumstances that do not meet the above criteria were regarded as an inadequate physical activity level.

### **Table S7.** Regression model for circulating metabolite and UPF intake in UKB (all 168 circulating metabolites)

| **UKB ID** | **Metabolite** | **Beta** | **FDR** | **Status** |
| --- | --- | --- | --- | --- |
| **Fatty acids** |  |  |  |  |
| 23442 | Total Fatty Acids | -0.037328 | 0.35677 | FDR<0.05& not Passed |
| 23444 | Omega-3 Fatty Acids | -0.39243 | 1.08E-24 | Passed elastic net model |
| 23445 | Omega-6 Fatty Acids | -0.150482 | 0.000145 | Passed elastic net model |
| 23446 | Polyunsaturated Fatty Acids | -0.241436 | 1.10E-09 | Passed elastic net model |
| 23447 | Monounsaturated Fatty Acids | 0.146609 | 0.000139 | FDR>0.05 |
| 23448 | Saturated Fatty Acids | -0.016739 | 0.6763 | Passed elastic net model |
| 23449 | Linoleic Acid | -0.14332 | 0.000265 | Passed elastic net model |
| 23450 | Docosahexaenoic Acid | -0.544197 | 1.01E-44 | Passed elastic net model |
| **Glycerolipids** |  |  |  |  |
| 23407 | Total Triglycerides | 0.187051 | 1.19E-06 | FDR>0.05 |
| 23408 | Triglycerides in VLDL | 0.210274 | 5.60E-08 | FDR>0.05 |
| 23409 | Triglycerides in LDL | 0.037873 | 0.344676 | FDR<0.05& not Passed |
| 23410 | Triglycerides in HDL | 0.088843 | 0.022682 | Passed elastic net model |
| 23436 | Total Cholines | -0.319551 | 5.34E-15 | FDR<0.05& not Passed |
| 23487 | Triglycerides in Chylomicrons and Extremely Large VLDL | 0.247409 | 2.59E-10 | FDR>0.05 |
| 23494 | Triglycerides in Very Large VLDL | 0.251173 | 1.24E-10 | FDR>0.05 |
| 23501 | Triglycerides in Large VLDL | 0.210007 | 5.88E-08 | FDR>0.05 |
| 23508 | Triglycerides in Medium VLDL | 0.159823 | 3.52E-05 | FDR>0.05 |
| 23515 | Triglycerides in Small VLDL | 0.151296 | 8.83E-05 | FDR>0.05 |
| 23522 | Triglycerides in Very Small VLDL | 0.080133 | 0.040584 | FDR<0.05& not Passed |
| 23529 | Triglycerides in IDL | 0.006083 | 0.871312 | Passed elastic net model |
| 23536 | Triglycerides in Large LDL | 0.006985 | 0.856975 | Passed elastic net model |
| 23543 | Triglycerides in Medium LDL | 0.069531 | 0.077622 | FDR<0.05& not Passed |
| 23550 | Triglycerides in Small LDL | 0.134797 | 0.000498 | FDR>0.05 |
| 23557 | Triglycerides in Very Large HDL | -0.102826 | 0.00893 | Passed elastic net model |
| 23564 | Triglycerides in Large HDL | -0.156881 | 8.05E-05 | Passed elastic net model |
| 23571 | Triglycerides in Medium HDL | 0.108585 | 0.005077 | FDR>0.05 |
| 23578 | Triglycerides in Small HDL | 0.239025 | 6.47E-10 | FDR>0.05 |
| **Glycerophospholipids** | |  |  |  |
| 23411 | Total Phospholipids in Lipoprotein Particles | -0.306735 | 3.31E-14 | Passed elastic net model |
| 23412 | Phospholipids in VLDL | 0.086929 | 0.025535 | Passed elastic net model |
| 23413 | Phospholipids in LDL | -0.191021 | 7.90E-07 | FDR<0.05& not Passed |
| 23414 | Phospholipids in HDL | -0.372862 | 2.47E-19 | Passed elastic net model |
| 23434 | Phosphoglycerides | -0.286466 | 1.98E-12 | Passed elastic net model |
| 23437 | Phosphatidylcholines | -0.321911 | 3.08E-15 | Passed elastic net model |
| 23483 | Phospholipids in Chylomicrons and Extremely Large VLDL | 0.257326 | 5.63E-11 | FDR>0.05 |
| 23490 | Phospholipids in Very Large VLDL | 0.222762 | 9.62E-09 | FDR>0.05 |
| 23497 | Phospholipids in Large VLDL | 0.217006 | 2.17E-08 | FDR>0.05 |
| 23504 | Phospholipids in Medium VLDL | -0.034203 | 0.389679 | Passed elastic net model |
| 23511 | Phospholipids in Small VLDL | -0.035318 | 0.37398 | Passed elastic net model |
| 23518 | Phospholipids in Very Small VLDL | -0.116757 | 0.003022 | Passed elastic net model |
| 23525 | Phospholipids in IDL | -0.316267 | 2.66E-15 | Passed elastic net model |
| 23532 | Phospholipids in Large LDL | -0.230329 | 3.42E-09 | FDR<0.05& not Passed |
| 23539 | Phospholipids in Medium LDL | -0.111282 | 0.004173 | FDR<0.05& not Passed |
| 23546 | Phospholipids in Small LDL | -0.175308 | 5.00E-06 | Passed elastic net model |
| 23553 | Phospholipids in Very Large HDL | -0.414129 | 4.44E-24 | FDR<0.05& not Passed |
| 23560 | Phospholipids in Large HDL | -0.446508 | 9.66E-27 | FDR<0.05& not Passed |
| 23567 | Phospholipids in Medium HDL | -0.26172 | 1.12E-10 | Passed elastic net model |
| 23574 | Phospholipids in Small HDL | -0.03422 | 0.389701 | Passed elastic net model |
| **Lipoprotein particles** | |  |  |  |
| 23423 | Total Lipids in Lipoprotein Particles | -0.201395 | 2.85E-07 | FDR<0.05& not Passed |
| 23424 | Total Lipids in VLDL | 0.130657 | 0.000712 | FDR>0.05 |
| 23425 | Total Lipids in LDL | -0.197562 | 3.37E-07 | FDR<0.05& not Passed |
| 23426 | Total Lipids in HDL | -0.408152 | 8.24E-23 | FDR<0.05& not Passed |
| 23427 | Total Concentration of Lipoprotein Particles | -0.329768 | 3.35E-16 | FDR<0.05& not Passed |
| 23428 | Concentration of VLDL Particles | 0.011444 | 0.767564 | Passed elastic net model |
| 23430 | Concentration of HDL Particles | -0.319884 | 2.10E-15 | FDR<0.05& not Passed |
| 23431 | Average Diameter for VLDL Particles | 0.357555 | 5.42E-19 | FDR>0.05 |
| 23432 | Average Diameter for LDL Particles | -0.256092 | 1.01E-10 | Passed elastic net model |
| 23433 | Average Diameter for HDL Particles | -0.443953 | 2.17E-26 | Passed elastic net model |
| 23439 | Apolipoprotein B | -0.147202 | 0.000142 | FDR<0.05& not Passed |
| 23440 | Apolipoprotein A1 | -0.364768 | 6.77E-19 | FDR<0.05& not Passed |
| 23464 | Total Concentration of Branched-Chain Amino Acids (Leucine + Isoleucine + Valine) | 0.044791 | 0.270531 | Passed elastic net model |
| 23481 | Concentration of Chylomicrons and Extremely Large VLDL Particles | 0.250031 | 1.54E-10 | FDR>0.05 |
| 23482 | Total Lipids in Chylomicrons and Extremely Large VLDL | 0.250976 | 1.41E-10 | FDR>0.05 |
| 23488 | Concentration of Very Large VLDL Particles | 0.233765 | 1.72E-09 | FDR>0.05 |
| 23489 | Total Lipids in Very Large VLDL | 0.232323 | 2.15E-09 | FDR>0.05 |
| 23495 | Concentration of Large VLDL Particles | 0.203941 | 1.38E-07 | FDR>0.05 |
| 23496 | Total Lipids in Large VLDL | 0.200171 | 2.33E-07 | FDR>0.05 |
| 23502 | Concentration of Medium VLDL Particles | -0.015542 | 0.689199 | FDR<0.05& not Passed |
| 23503 | Total Lipids in Medium VLDL | 0.029157 | 0.461446 | FDR<0.05& not Passed |
| 23509 | Concentration of Small VLDL Particles | 0.056797 | 0.150877 | FDR<0.05& not Passed |
| 23510 | Total Lipids in Small VLDL | 0.041137 | 0.302487 | FDR<0.05& not Passed |
| 23516 | Concentration of Very Small VLDL Particles | -0.151228 | 0.000113 | Passed elastic net model |
| 23517 | Total Lipids in Very Small VLDL | -0.151708 | 0.000113 | Passed elastic net model |
| 23523 | Concentration of IDL Particles | -0.19343 | 7.58E-07 | Passed elastic net model |
| 23524 | Total Lipids in IDL | -0.313373 | 3.89E-15 | Passed elastic net model |
| 23530 | Concentration of Large LDL Particles | -0.180827 | 3.05E-06 | FDR<0.05& not Passed |
| 23531 | Total Lipids in Large LDL | -0.246236 | 2.91E-10 | FDR<0.05& not Passed |
| 23537 | Concentration of Medium LDL Particles | -0.100438 | 0.009642 | Passed elastic net model |
| 23538 | Total Lipids in Medium LDL | -0.106625 | 0.006146 | Passed elastic net model |
| 23544 | Concentration of Small LDL Particles | -0.105171 | 0.006706 | Passed elastic net model |
| 23545 | Total Lipids in Small LDL | -0.127052 | 0.001003 | FDR<0.05& not Passed |
| 23551 | Concentration of Very Large HDL Particles | -0.437357 | 1.11E-26 | Passed elastic net model |
| 23552 | Total Lipids in Very Large HDL | -0.427659 | 1.29E-25 | FDR<0.05& not Passed |
| 23558 | Concentration of Large HDL Particles | -0.466531 | 1.14E-28 | FDR<0.05& not Passed |
| 23559 | Total Lipids in Large HDL | -0.4578 | 5.52E-28 | FDR<0.05& not Passed |
| 23565 | Concentration of Medium HDL Particles | -0.334599 | 2.84E-16 | Passed elastic net model |
| 23566 | Total Lipids in Medium HDL | -0.299942 | 1.58E-13 | Passed elastic net model |
| 23572 | Concentration of Small HDL Particles | -0.065884 | 0.096539 | Passed elastic net model |
| 23573 | Total Lipids in Small HDL | -0.032528 | 0.411606 | Passed elastic net model |
| **Sphingolipids** | |  |  |  |
| 23438 | Sphingomyelins | -0.370752 | 1.16E-19 | FDR<0.05& not Passed |
| **Sterol lipids** |  |  |  |  |
| 23400 | Total Cholesterol | -0.322801 | 6.08E-16 | Passed elastic net model |
| 23401 | Total Cholesterol Minus HDL-C | -0.196407 | 4.37E-07 | Passed elastic net model |
| 23402 | Remnant Cholesterol (Non-HDL, Non-LDL -Cholesterol) | -0.172326 | 9.91E-06 | Passed elastic net model |
| 23403 | VLDL Cholesterol | -0.016403 | 0.6763 | Passed elastic net model |
| 23404 | Clinical LDL Cholesterol | -0.221282 | 1.31E-08 | FDR<0.05& not Passed |
| 23405 | LDL Cholesterol | -0.21218 | 4.71E-08 | Passed elastic net model |
| 23406 | HDL Cholesterol | -0.447388 | 4.03E-27 | FDR<0.05& not Passed |
| 23415 | Total Esterified Cholesterol | -0.338378 | 2.85E-17 | FDR<0.05& not Passed |
| 23416 | Cholesteryl Esters in VLDL | -0.065518 | 0.098198 | Passed elastic net model |
| 23417 | Cholesteryl Esters in LDL | -0.189015 | 1.06E-06 | FDR<0.05& not Passed |
| 23418 | Cholesteryl Esters in HDL | -0.440112 | 1.52E-26 | FDR<0.05& not Passed |
| 23419 | Total Free Cholesterol | -0.277331 | 2.71E-12 | FDR<0.05& not Passed |
| 23420 | Free Cholesterol in VLDL | 0.050134 | 0.204044 | Passed elastic net model |
| 23421 | Free Cholesterol in LDL | -0.267425 | 8.31E-12 | FDR<0.05& not Passed |
| 23422 | Free Cholesterol in HDL | -0.456339 | 1.03E-27 | FDR<0.05& not Passed |
| 23429 | Concentration of LDL Particles | -0.153696 | 7.29E-05 | FDR<0.05& not Passed |
| 23484 | Cholesterol in Chylomicrons and Extremely Large VLDL | 0.245879 | 2.91E-10 | FDR>0.05 |
| 23485 | Cholesteryl Esters in Chylomicrons and Extremely Large VLDL | 0.243811 | 3.77E-10 | FDR>0.05 |
| 23486 | Free Cholesterol in Chylomicrons and Extremely Large VLDL | 0.24432 | 3.77E-10 | FDR>0.05 |
| 23491 | Cholesterol in Very Large VLDL | 0.169853 | 1.08E-05 | FDR>0.05 |
| 23492 | Cholesteryl Esters in Very Large VLDL | 0.131073 | 0.000682 | FDR>0.05 |
| 23493 | Free Cholesterol in Very Large VLDL | 0.204854 | 1.22E-07 | FDR>0.05 |
| 23498 | Cholesterol in Large VLDL | 0.152858 | 7.47E-05 | FDR>0.05 |
| 23499 | Cholesteryl Esters in Large VLDL | 0.100613 | 0.009356 | Passed elastic net model |
| 23500 | Free Cholesterol in Large VLDL | 0.200144 | 2.33E-07 | FDR>0.05 |
| 23505 | Cholesterol in Medium VLDL | -0.169279 | 1.29E-05 | FDR<0.05& not Passed |
| 23506 | Cholesteryl Esters in Medium VLDL | -0.231903 | 2.93E-09 | FDR<0.05& not Passed |
| 23507 | Free Cholesterol in Medium VLDL | -0.078724 | 0.044648 | Passed elastic net model |
| 23512 | Cholesterol in Small VLDL | -0.050676 | 0.199985 | FDR<0.05& not Passed |
| 23513 | Cholesteryl Esters in Small VLDL | -0.02171 | 0.587388 | FDR<0.05& not Passed |
| 23514 | Free Cholesterol in Small VLDL | -0.099786 | 0.010227 | Passed elastic net model |
| 23519 | Cholesterol in Very Small VLDL | -0.243515 | 9.82E-10 | FDR<0.05& not Passed |
| 23520 | Cholesteryl Esters in Very Small VLDL | -0.27343 | 9.30E-12 | Passed elastic net model |
| 23521 | Free Cholesterol in Very Small VLDL | -0.163868 | 3.19E-05 | Passed elastic net model |
| 23526 | Cholesterol in IDL | -0.325693 | 3.42E-16 | FDR<0.05& not Passed |
| 23527 | Cholesteryl Esters in IDL | -0.322502 | 6.42E-16 | FDR<0.05& not Passed |
| 23528 | Free Cholesterol in IDL | -0.32548 | 3.22E-16 | Passed elastic net model |
| 23533 | Cholesterol in Large LDL | -0.261452 | 2.53E-11 | FDR<0.05& not Passed |
| 23534 | Cholesteryl Esters in Large LDL | -0.245205 | 3.29E-10 | Passed elastic net model |
| 23535 | Free Cholesterol in Large LDL | -0.299013 | 2.91E-14 | Passed elastic net model |
| 23540 | Cholesterol in Medium LDL | -0.115526 | 0.002977 | Passed elastic net model |
| 23541 | Cholesteryl Esters in Medium LDL | -0.079692 | 0.042479 | Passed elastic net model |
| 23542 | Free Cholesterol in Medium LDL | -0.205561 | 1.07E-07 | FDR<0.05& not Passed |
| 23547 | Cholesterol in Small LDL | -0.128855 | 0.000871 | Passed elastic net model |
| 23548 | Cholesteryl Esters in Small LDL | -0.092321 | 0.018101 | Passed elastic net model |
| 23549 | Free Cholesterol in Small LDL | -0.212843 | 3.56E-08 | Passed elastic net model |
| 23554 | Cholesterol in Very Large HDL | -0.440459 | 4.03E-27 | FDR<0.05& not Passed |
| 23555 | Cholesteryl Esters in Very Large HDL | -0.452029 | 3.78E-28 | Passed elastic net model |
| 23556 | Free Cholesterol in Very Large HDL | -0.380863 | 2.11E-21 | FDR<0.05& not Passed |
| 23561 | Cholesterol in Large HDL | -0.463131 | 1.14E-28 | FDR<0.05& not Passed |
| 23562 | Cholesteryl Esters in Large HDL | -0.462097 | 1.14E-28 | FDR<0.05& not Passed |
| 23563 | Free Cholesterol in Large HDL | -0.461997 | 1.99E-28 | Passed elastic net model |
| 23568 | Cholesterol in Medium HDL | -0.352686 | 5.16E-18 | FDR<0.05& not Passed |
| 23569 | Cholesteryl Esters in Medium HDL | -0.344023 | 2.65E-17 | FDR<0.05& not Passed |
| 23570 | Free Cholesterol in Medium HDL | -0.37828 | 4.51E-20 | FDR<0.05& not Passed |
| 23575 | Cholesterol in Small HDL | -0.09511 | 0.014728 | Passed elastic net model |
| 23576 | Cholesteryl Esters in Small HDL | -0.062658 | 0.113135 | Passed elastic net model |
| 23577 | Free Cholesterol in Small HDL | -0.181419 | 3.32E-06 | Passed elastic net model |
| **Amino acids** |  |  |  |  |
| 23460 | Alanine | 0.039601 | 0.314423 | FDR<0.05& not Passed |
| 23461 | Glutamine | 0.021372 | 0.588365 | Passed elastic net model |
| 23462 | Glycine | 0.059171 | 0.155866 | Passed elastic net model |
| 23463 | Histidine | -0.054316 | 0.171472 | FDR<0.05& not Passed |
| 23465 | Isoleucine | 0.06071 | 0.127921 | Passed elastic net model |
| 23466 | Leucine | 0.07235 | 0.072613 | FDR<0.05& not Passed |
| 23467 | Valine | 0.016864 | 0.6763 | Passed elastic net model |
| 23468 | Phenylalanine | 0.051276 | 0.198612 | FDR<0.05& not Passed |
| 23469 | Tyrosine | -0.022734 | 0.57348 | Passed elastic net model |
| **Carbohydrates and Derivatives** | |  |  |  |
| 23470 | Glucose | 0.15187 | 0.000109 | FDR>0.05 |
| 23471 | Lactate | 0.226244 | 4.48E-09 | FDR>0.05 |
| 23472 | Pyruvate | 0.249212 | 1.37E-10 | FDR>0.05 |
| 23473 | Citrate | 0.036768 | 0.362221 | Passed elastic net model |
| 23474 | 3-Hydroxybutyrate | -0.063637 | 0.110243 | FDR<0.05& not Passed |
| 23475 | Acetate | -0.18443 | 2.24E-06 | Passed elastic net model |
| 23476 | Acetoacetate | -0.054617 | 0.171631 | FDR<0.05& not Passed |
| 23477 | Acetone | -0.137734 | 0.000304 | Passed elastic net model |
| **Fluid Balance and inflammation** | |  |  |  |
| 23478 | Creatinine | 0.269672 | 2.61E-09 | FDR>0.05 |
| 23479 | Albumin | -0.207026 | 1.07E-07 | Passed elastic net model |
| 23480 | Glycoprotein Acetyls | 0.334692 | 4.45E-18 | FDR>0.05 |

### **Table S8.** Associations between UPF intake and its metabolic signature and incident IBD in UK Biobank ^a^

| **UPF intake** | IBD |  |  | CD |  |  | UC |  |  |
| --- | --- | --- | --- | --- | --- | --- | --- | --- | --- |
|  | Cases | HR (95% CI) | *P* | Cases | HR (95% CI) | *P* | Cases | HR (95% CI) | *P* |
| **UPF intake** |  |  |  |  |  |  |  |  |  |
| **In Serving/day** |  |  |  |  |  |  |  |  |  |
| Per SD | 983 | **1.08 (1.01, 1.15)** | **0.017** | 295 | **1.14 (1.02, 1.28)** | **0.024** | 645 | 1.03 (0.95, 1.12) | 0.444 |
| T1 (lowest) | 293 | Ref |  | 83 | Ref |  | 199 | Ref |  |
| T2 | 312 | 1.04 (0.88, 1.22) | 0.657 | 84 | 1.02 (0.74, 1.38) | 0.925 | 217 | 1.05 (0.86, 1.27) | 0.657 |
| T3 (highest) | 378 | **1.21 (1.02, 1.43)** | **0.032** | 128 | **1.52 (1.11, 2.06)** | **0.008** | 229 | 1.04 (0.84, 1.29) | 0.704 |
| *P* for trend |  |  | **0.019** |  |  | **0.006** |  |  | 0.605 |
| **In Energy/day** |  |  |  |  |  |  |  |  |  |
| Per SD |  | **1.12 (1.04, 1.22)** | **0.005** |  | **1.26 (1.09, 1.46)** | **0.002** |  | 1.04 (0.94, 1.15) | 0.503 |
| T1 (lowest) | 293 | Ref |  | 82 | Ref |  | 202 | Ref |  |
| T2 | 327 | 1.12 (0.95, 1.32) | 0.166 | 89 | 1.14 (0.84, 1.56) | 0.398 | 223 | 1.08 (0.89, 1.32) | 0.432 |
| T3 (highest) | 363 | 1.20 (0.99, 1.45) | 0.060 | 124 | **1.61 (1.14, 2.28)** | **0.007** | 220 | 0.99 (0.79, 1.26) | 0.966 |
| *P* for trend |  |  | 0.059 |  |  | **0.006** |  |  | 0.988 |
| **In Energy proportion/day** | |  |  |  |  |  |  |  |  |
| Per SD |  | **1.11 (1.04, 1.18)** | **0.001** |  | **1.24 (1.11, 1.39)** | **<0.001** |  | 1.04 (0.96, 1.12) | 0.363 |
| T1 (lowest) | 295 | Ref |  | 74 | Ref |  | 211 | Ref |  |
| T2 | 324 | 1.11 (0.95, 1.30) | 0.198 | 106 | **1.45 (1.08, 1.96)** | **0.015** | 207 | 0.99 (0.82, 1.20) | 0.922 |
| T3 (highest) | 364 | **1.22 (1.04, 1.42)** | **0.015** | 115 | **1.51 (1.12, 2.03)** | **0.007** | 227 | 1.06 (0.88, 1.29) | 0.524 |
| *P* for trend |  |  | **0.014** |  |  | **0.009** |  |  | 0.519 |
| **UPF metabolic score** | | | | | | | | | |
| **Discovery cohort (n=10,229)** | | | | | | | | | |
| Per SD |  | **1.71 (1.19, 2.46)** | **0.004** |  | **2.51 (1.16, 5.46)** | **0.02** |  | 1.50 (0.99, 2.27) | 0.055 |
| Low | 13 | Ref |  | 1 | Ref |  | 12 | Ref |  |
| High | 35 | **2.20 (1.09, 4.45)** | **0.028** | 11 | 4.68 (0.57, 38.71) | 0.152 | 23 | 1.93 (0.89, 4.22) | 0.097 |
| **Internal validation cohort (n=91,306)** | | | | | | | | | |
| Per SD |  | **1.24 (1.11, 1.38)** | **<0.001** |  | **1.46 (1.25, 1.70)** | **<0.001** |  | 1.13 (0.99, 1.29) | 0.073 |
| T1 (lowest) | 111 | Ref |  | 28 | Ref |  | 78 | Ref |  |
| T2 | 178 | **1.58 (1.23, 2.02)** | **<0.001** | 44 | **1.68 (1.03, 2.73)** | **0.038** | 128 | **1.56 (1.17, 2.10)** | **0.003** |
| T3 (highest) | 192 | **1.65 (1.25, 2.18)** | **<0.001** | 63 | **2.65 (1.57, 4.48)** | **<0.001** | 119 | 1.36 (0.97, 1.90) | 0.078 |
| *P* for trend |  |  | **0.001** |  |  | **<0.001** |  |  | 0.106 |

^a^ Adjusted for age, sex, Townsend deprivation index, physical activity, smoking status, BMI, total energy, and total sugar intake.

CD, Crohn’s disease; CI, confidence interval; HR, hazard ratio; IBD, inflammatory bowel disease; SD, standard deviation; T1 to T3, tertile 1 to tertile 3; UC, ulcerative colitis; UPF, ultra-processed food.

### **Table S9.** Joint association and additive interaction of UPF, UPF metabolic score and genetic risk and incident CD

| Exposure subgroups | Incident CD |  |  | Additive interaction | |  |  |  |  |
| --- | --- | --- | --- | --- | --- | --- | --- | --- | --- |
|  | Case/person-years | HR (95% CI) ^a^ | *P* | RERI (95% CI) | *P* | AP (95% CI) | *P* | SI (95% CI) | *P* |
| **UPF intake** |  |  |  | **0.62 (0.02, 1.22)** | **0.021** | **0.29 (0.02, 0.57)** | **0.019** | 2.22 (0.64, 7.69) | 0.103 |
| low UPF, low PRS | 51/496,152 | Ref |  |  |  |  |  |  |  |
| low UPF, high PRS | 65/501,379 | 1.26 (0.88, 1.82) | 0.213 |  |  |  |  |  |  |
| high UPF, low PRS | 64/498,333 | 1.21 (0.82, 1.77) | 0.336 |  |  |  |  |  |  |
| high UPF, high PRS | 108/492,190 | **2.06 (1.46, 2.92)** | **<0.001** |  |  |  |  |  |  |
| **UPF metabolic score** |  |  |  | **0.40 (0.08, 0.72)** | **0.007** | 0.28 (-0.12, 0.67) | 0.085 | 14.83 (0.01, 146.12) | 0.456 |
| low UPF metabolic score, low PRS | 25/246,694 | Ref |  |  |  |  |  |  |  |
| low UPF metabolic score, high PRS | 29/247,876 | 1.17 (0.69, 2.00) | 0.563 |  |  |  |  |  |  |
| high UPF metabolic score, low PRS | 30/246,171 | 1.21 (0.69, 2.14) | 0.503 |  |  |  |  |  |  |
| high UPF metabolic score, high PRS | 51/244,988 | **2.07 (1.23, 3.47)** | **0.006** |  |  |  |  |  |  |

^a^ Based on the fully-adjusted model adjusted for age, sex, Townsend deprivation index, physical activity, smoking status, BMI, total energy, and total sugar intake.

AP, attributable proportion; CD, Crohn’s disease; CI, confidence interval; HR, hazard ratio; PRS, polygenic risk score; RERI, relative excess risk due to interaction; SI, synergy index; UPF, ultra-processed food.

### **Table S10.** Modules of metabolites filter through the elastic net model, categorized by WGCNA ^a^

| Category of metabolites | Turquoise | Blue | Yellow | Brown | Grey (unclassified) |
| --- | --- | --- | --- | --- | --- |
| **Total metabolites** | 31 | 20 | 4 | 10 | 8 |
| Sterol lipids | 16 | 6 | 2 | 2 | 0 |
| Fatty acids | 5 | 1 | 0 | 0 | 0 |
| Glycerolipids | 0 | 7 | 0 | 1 | 0 |
| Glycerophospholipids | 3 | 2 | 0 | 5 | 0 |
| Lipoprotein particles | 7 | 3 | 2 | 2 | 1 |
| Carbohydrates and derivatives | 0 | 0 | 0 | 0 | 5 |
| Fluid balance and inflammation | 0 | 1 | 0 | 0 | 2 |

^a^ WGCNA, weighted co-expression network analysis.

### **Table S11.** Association between the combined z-score of metabolites in metabolite modules and incident IBD and CD ^a^

| Module | IBD |  | CD |  |
| --- | --- | --- | --- | --- |
|  | HR (95% CI) | *P* | HR (95% CI) | *P* |
| Blue | **1.01 (0.95, 1.08)** | **0.001** | 0.93 (0.83, 1.04) | 0.196 |
| Yellow | 1.01 (0.95, 1.07) | 0.059 | 0.93 (0.96, 7.95) | 0.193 |
| Brown | 0.99 (0.93, 1.05) | 0.680 | 0.90 (0.80, 1.00) | 0.059 |
| Turquoise | 0.98 (0.93, 1.05) | 0.601 | **0.87 (0.77, 0.98)** | **0.019** |

^a^ Adjusted for age, sex, Townsend deprivation index, physical activity, smoking status, BMI, total energy, and total sugar intake.

CD, Crohn’s disease; CI, confidence interval; HR, hazard ratio; IBD, inflammatory bowel disease; SD, standard deviation.

### **Table S12.** The coefficient derived from the elastic net regression and WGCNA module eigengenes of metabolites on incident IBD and CD.

| UKB ID | Metabolite | Coefficient in the elastic net regression | Module eigengenes ^a^ | |
| --- | --- | --- | --- | --- |
|  |  |  | IBD | CD |
| **Turquoise** |  |  |  |  |
| 23400 | Total Cholesterol | 0.33139 | -0.02836 | -0.026563 |
| 23401 | Total Cholesterol Minus HDL-C | 0.08614 | -0.021176 | -0.017495 |
| 23402 | Remnant Cholesterol (Non-HDL, Non-LDL -Cholesterol) | 0.02327 | -0.018195 | -0.010777 |
| 23405 | LDL Cholesterol | 0.01693 | -0.023297 | -0.023258 |
| 23444 | Omega-3 Fatty Acids | 0.53039 | -0.018443 | -0.002878 |
| 23445 | Omega-6 Fatty Acids | 1.51591 | -0.017474 | -0.012382 |
| 23446 | Polyunsaturated Fatty Acids | 0.01521 | -0.01997 | -0.011349 |
| 23449 | Linoleic Acid | -0.68429 | -0.01527 | -0.01135 |
| 23450 | Docosahexaenoic Acid | -0.77282 | -0.025657 | -0.017452 |
| 23507 | Free Cholesterol in Medium VLDL | 0.03825 | -0.008799 | -0.00343 |
| 23514 | Free Cholesterol in Small VLDL | 0.13822 | -0.012192 | -0.006467 |
| 23516 | Concentration of Very Small VLDL Particles | -0.02328 | -0.018666 | -0.005062 |
| 23517 | Total Lipids in Very Small VLDL | 0.23643 | -0.018264 | -0.003641 |
| 23518 | Phospholipids in Very Small VLDL | 0.88897 | -0.015197 | -3.33E-07 |
| 23520 | Cholesteryl Esters in Very Small VLDL | -0.25101 | -0.028462 | -0.017609 |
| 23521 | Free Cholesterol in Very Small VLDL | 0.65602 | -0.019514 | -0.006571 |
| 23523 | Concentration of IDL Particles | 0.71606 | -0.025367 | -0.018725 |
| 23524 | Total Lipids in IDL | -1.27609 | -0.029491 | -0.022716 |
| 23525 | Phospholipids in IDL | -0.56392 | -0.031141 | -0.024632 |
| 23528 | Free Cholesterol in IDL | 0.22282 | -0.030982 | -0.026694 |
| 23534 | Cholesteryl Esters in Large LDL | 0.17209 | -0.025589 | -0.025479 |
| 23535 | Free Cholesterol in Large LDL | -0.31391 | -0.029825 | -0.030458 |
| 23537 | Concentration of Medium LDL Particles | -0.50350 | -0.013288 | -0.011794 |
| 23538 | Total Lipids in Medium LDL | -0.02178 | -0.014518 | -0.013515 |
| 23540 | Cholesterol in Medium LDL | -0.00201 | -0.015349 | -0.015071 |
| 23541 | Cholesteryl Esters in Medium LDL | -0.00037 | -0.011667 | -0.010537 |
| 23544 | Concentration of Small LDL Particles | -0.55654 | -0.012689 | -0.010466 |
| 23546 | Phospholipids in Small LDL | -1.04029 | -0.019141 | -0.014979 |
| 23547 | Cholesterol in Small LDL | 0.77743 | -0.016503 | -0.016245 |
| 23548 | Cholesteryl Esters in Small LDL | 0.01696 | -0.012751 | -0.011387 |
| 23549 | Free Cholesterol in Small LDL | 0.01396 | -0.024687 | -0.027339 |
| **Blue** |  |  |  |  |
| 23407 | Total Triglycerides | 0.195615 | 0.0202998 | 0.0286391 |
| 23408 | Triglycerides in VLDL | 0.426445 | 0.0230322 | 0.0298863 |
| 23412 | Phospholipids in VLDL | 0.055636 | 0.0072198 | 0.0158949 |
| 23431 | Average Diameter for VLDL Particles | 0.139543 | 0.0332121 | 0.0320619 |
| 23447 | Monounsaturated Fatty Acids | -0.466437 | 0.0081139 | 0.01467 |
| 23480 | Glycoprotein Acetyls | 0.241004 | 0.0156079 | 0.0194411 |
| 23481 | Concentration of Chylomicrons and Extremely Large VLDL Particles | 0.214073 | 0.0224763 | 0.0292293 |
| 23485 | Cholesteryl Esters in Chylomicrons and Extremely Large VLDL | -0.239215 | 0.0213967 | 0.0221565 |
| 23486 | Free Cholesterol in Chylomicrons and Extremely Large VLDL | 0.46667 | 0.0210176 | 0.0273921 |
| 23493 | Free Cholesterol in Very Large VLDL | -0.700121 | 0.0194922 | 0.0265469 |
| 23495 | Concentration of Large VLDL Particles | -0.001108 | 0.0218584 | 0.0275575 |
| 23497 | Phospholipids in Large VLDL | -1.057415 | 0.0213902 | 0.0281766 |
| 23498 | Cholesterol in Large VLDL | -0.043126 | 0.0156024 | 0.0223201 |
| 23499 | Cholesteryl Esters in Large VLDL | -0.734194 | 0.010285 | 0.0171017 |
| 23500 | Free Cholesterol in Large VLDL | -1.068927 | 0.0202062 | 0.026644 |
| 23501 | Triglycerides in Large VLDL | 0.718019 | 0.0253439 | 0.0308683 |
| 23508 | Triglycerides in Medium VLDL | 1.054152 | 0.0205343 | 0.025655 |
| 23515 | Triglycerides in Small VLDL | -0.40236 | 0.0183731 | 0.027071 |
| 23522 | Triglycerides in Very Small VLDL | 0.01622 | 0.0059768 | 0.0213542 |
| 23557 | Triglycerides in Very Large HDL | -0.604313 | -0.004814 | 0.0053574 |
| **Yellow** |  |  |  |  |
| 23433 | Average Diameter for HDL Particles | -0.27913 | -0.029208 | -0.028545 |
| 23551 | Concentration of Very Large HDL Particles | 0.672531 | -0.027986 | -0.028954 |
| 23555 | Cholesteryl Esters in Very Large HDL | 1.11E-05 | -0.029095 | -0.031376 |
| 23563 | Free Cholesterol in Large HDL | 0.009351 | -0.028753 | -0.030094 |
| **Brown** |  |  |  |  |
| 23411 | Total Phospholipids in Lipoprotein Particles | -0.05688 | -0.023675 | -0.01816 |
| 23414 | Phospholipids in HDL | -0.26871 | -0.023228 | -0.021694 |
| 23434 | Phosphoglycerides | 0.436423 | -0.021833 | -0.016106 |
| 23437 | Phosphatidylcholines | -0.01084 | -0.022323 | -0.016589 |
| 23564 | Triglycerides in Large HDL | 0.096799 | -0.007763 | 0.002909 |
| 23565 | Concentration of Medium HDL Particles | -0.53628 | -0.021976 | -0.022967 |
| 23566 | Total Lipids in Medium HDL | -0.65223 | -0.019688 | -0.019081 |
| 23567 | Phospholipids in Medium HDL | -0.58676 | -0.016932 | -0.013994 |
| 23575 | Cholesterol in Small HDL | 0.078211 | -0.010746 | -0.014564 |
| 23577 | Free Cholesterol in Small HDL | 1.056863 | -0.014882 | -0.012971 |
| **Grey (unclassified)** | |  |  |  |
| 23432 | Average Diameter for LDL Particles | -0.17960 | -0.024003 | -0.022514 |
| 23470 | Glucose | 0.065681 | -0.00155 | 0.0057121 |
| 23471 | Lactate | 0.106023 | 0.0109694 | 0.0118561 |
| 23472 | Pyruvate | -0.22896 | 0.012301 | 0.0118118 |
| 23475 | Acetate | -0.02961 | -0.008574 | -0.005411 |
| 23477 | Acetone | -0.14213 | -0.001644 | 0.0062955 |
| 23478 | Creatinine | 0.378408 | 0.0226263 | 0.0306971 |
| 23479 | Albumin | -0.06213 | -0.01306 | -0.000639 |

^a^ The eigengenes representing the Pearson correlation coefficients between single metabolites in WGCNA modules and incident IBD and CD.

CD, Crohn’s disease; IBD, inflammatory bowel disease; SD, standard deviation; UC, ulcerative colitis; UPF, ultra-processed food; WGCNA, weighted co-expression network analysis.

### **Table S13.** Mediation analysis of metabolites that show significant associations with incident CD (ranked by mediating proportion).

| Metabolites | ACME (E-08) | 95% CI (E-08) | *P* | ADE (E-07) | 95% CI (E-07) | *P* | Mediating proportion (%) | 95% CI | *P* |
| --- | --- | --- | --- | --- | --- | --- | --- | --- | --- |
| DHA | 16.46 | (7.91, 24.47) | <1e-16 | 7.15 | (1.50, 13.73) | <1e-16 | 17.14 | (5.96, 51.95) | <1e-16 |
| Free Cholesterol in IDL | 14.21 | (7.68, 20.17) | <1e-16 | 7.40 | (1.38, 13.62) | <1e-16 | 15.52 | (6.89, 46.95) | <1e-16 |
| Total Cholesterol | 14.18 | (8.54, 19.96) | <1e-16 | 7.81 | (2.20, 13.31) | <1e-16 | 14.84 | (7.36, 40.67) | <1e-16 |
| Total Lipids in IDL | 12.96 | (6.62, 18.11) | <1e-16 | 7.73 | (2.15, 13.32) | <1e-16 | 13.82 | (6.71, 42.07) | <1e-16 |
| Phosphatidylcholines | 12.73 | (6.02, 19.01) | <1e-16 | 7.81 | (2.05, 13.34) | <1e-16 | 13.36 | (5.84, 42.13) | <1e-16 |
| Free Cholesterol in Large LDL | 12.31 | (6.49, 17.54) | <1e-16 | 7.63 | (1.51, 13.52) | <1e-16 | 13.29 | (6.08, 42.14) | <1e-16 |
| Total Phospholipids in Lipoprotein Particles | 12.39 | (6.77, 17.95) | <1e-16 | 7.94 | (2.29, 13.54) | <1e-16 | 13.23 | (6.12, 38.08) | <1e-16 |
| Concentration of Medium HDL Particles | 10.53 | (3.03, 17.52) | <1e-16 | 7.71 | (2.46, 14.28) | <1e-16 | 12.51 | (2.73, 35.69) | <1e-16 |
| Phospholipids in HDL | 11.25 | (2.75, 18.63) | <1e-16 | 7.85 | (1.72, 13.60) | <1e-16 | 12.46 | (3.07, 44.71) | <1e-16 |
| Phospholipids in IDL | 11.38 | (5.57, 16.69) | <1e-16 | 7.70 | (1.56, 13.78) | <1e-16 | 12.3 | (4.90, 37.98) | <1e-16 |
| Phosphoglycerides | 11.69 | (5.81, 17.27) | <1e-16 | 7.95 | (2.22, 13.47) | <1e-16 | 12.14 | (5.62, 36.45) | <1e-16 |
| Free Cholesterol in Large HDL | 10.24 | (0.84, 19.00) | 0.040 | 7.74 | (2.40, 14.43) | <1e-16 | 11.92 | (0.80, 38.83) | 0.040 |
| Omega-3 Fatty Acids | 11.08 | (4.88, 16.71) | <1e-16 | 7.73 | (1.57, 13.92) | <1e-16 | 11.79 | (4.25, 39.29) | <1e-16 |
| Concentration of Very Large HDL Particles | 9.95 | (0.82, 18.35) | 0.040 | 7.77 | (2.47, 14.49) | <1e-16 | 11.21 | (0.80, 38.09) | 0.040 |
| Total Lipids in Medium HDL | 9.49 | (1.75, 16.01) | <1e-16 | 7.98 | (1.73, 13.74) | <1e-16 | 10.22 | (1.86, 37.45) | <1e-16 |
| Cholesteryl Esters in Very Small VLDL | 8.72 | (3.94, 13.25) | <1e-16 | 7.92 | (2.27, 14.44) | <1e-16 | 9.36 | (3.32, 26.22) | <1e-16 |
| Cholesteryl Esters in Large LDL | 8.14 | (4.04, 11.76) | <1e-16 | 8.18 | (2.63, 13.73) | <1e-16 | 8.87 | (4.10, 25.99) | <1e-16 |
| Polyunsaturated Fatty Acids | 8.15 | (3.89, 12.58) | <1e-16 | 8.43 | (2.74, 13.99) | <1e-16 | 8.31 | (3.48, 24.79) | <1e-16 |
| Phospholipids in Medium HDL | 7.75 | (1.06, 13.74) | <1e-16 | 8.06 | (1.96, 14.29) | <1e-16 | 7.88 | (0.89, 27.33) | <1e-16 |
| Concentration of IDL Particles | 6.41 | (3.26, 9.81) | <1e-16 | 8.14 | (2.51, 14.65) | <1e-16 | 7.09 | (2.89, 18.63) | <1e-16 |
| Free Cholesterol in Small HDL | 6.35 | (2.71, 9.93) | <1e-16 | 8.17 | (2.30, 14.59) | <1e-16 | 6.92 | (2.43, 18.59) | <1e-16 |
| LDL-Cholesterol | 6.41 | (3.18, 9.13) | <1e-16 | 8.47 | (2.95, 14.00) | <1e-16 | 6.78 | (3.37, 18.41) | <1e-16 |
| Free Cholesterol in Small LDL | 5.73 | (2.47, 9.15) | <1e-16 | 8.23 | (2.26, 14.45) | <1e-16 | 6.33 | (2.37, 16.16) | <1e-16 |
| Albumin | 5.96 | (3.10, 8.84) | <1e-16 | 8.77 | (2.14, 13.54) | <1e-16 | 6.05 | (2.62, 21.04) | <1e-16 |
| "Total Cholesterol |  |  |  |  |  |  |  |  |  |
| minus HDL-C" | 5.54 | (2.94, 8.14) | <1e-16 | 8.62 | (3.02, 13.98) | <1e-16 | 5.76 | (2.95, 16.67) | <1e-16 |
| Omega-6 Fatty Acids | 5.01 | (1.97, 8.20) | 0.020 | 8.76 | (3.10, 14.23) | <1e-16 | 5.19 | (1.88, 15.48) | 0.020 |
| Remnant Cholesterol (Non-HDL, Non-LDL -Cholesterol) | 4.22 | (1.99, 6.21) | <1e-16 | 8.63 | (3.05, 14.10) | <1e-16 | 4.52 | (2.11, 12.28) | <1e-16 |
| Cholesterol in Small HDL | 3.85 | (1.16, 6.44) | <1e-16 | 8.52 | (2.46, 14.07) | <1e-16 | 4.27 | (1.02, 12.78) | <1e-16 |

ACME, average causal mediation effect; ADE, average direct effect; CD, Crohn’s disease; CI, confidence interval; DHA, docosahexaenoic acid; HDL, high-density lipoprotein; IDL, intermediate density lipoprotein; LDL, low-density lipoprotein.

### **Table S14.** Metabolites in the metabolic signature with a significant association with incident IBD and CD

| UKB ID | Metabolite | IBD | | CD | |
| --- | --- | --- | --- | --- | --- |
|  |  | HR (95% CI) ^a^ | *P* | HR (95% CI) | *P* |
| 23400 | Total Cholesterol | 0.84 (0.76, 0.92) | <0.001 | 0.60 (0.49, 0.73) | <0.001 |
| 23401 | Total Cholesterol Minus HDL-C | 0.87 (0.79, 0.97) | 0.012 | 0.61 (0.49, 0.76) | <0.001 |
| 23402 | Remnant Cholesterol (Non-HDL, Non-LDL -Cholesterol) | 0.79 (0.64, 0.98) | 0.029 | 0.38 (0.24, 0.60) | <0.001 |
| 23405 | LDL Cholesterol | 0.76 (0.62, 0.92) | 0.006 | 0.39 (0.26, 0.60) | <0.001 |
| 23411 | Total Phospholipids in Lipoprotein Particles | 0.70 (0.58, 0.86) | <0.001 | 0.38 (0.25, 0.56) | <0.001 |
| 23433 | Average Diameter for HDL Particles | 0.48 (0.29, 0.82) | 0.007 | 0.50 (0.18, 1.41) | 0.193 |
| 23434 | Phosphoglycerides | 0.66 (0.53, 0.84) | 0.001 | 0.33 (0.21, 0.54) | <0.001 |
| 23437 | Phosphatidylcholines | 0.63 (0.49, 0.81) | <0.001 | 0.32 (0.19, 0.53) | <0.001 |
| 23444 | Omega-3 Fatty Acids | 0.62 (0.41, 0.94) | 0.025 | 0.16 (0.06, 0.42) | <0.001 |
| 23445 | Omega-6 Fatty Acids | 0.86 (0.76, 0.98) | 0.027 | 0.64 (0.49, 0.84) | 0.001 |
| 23446 | Polyunsaturated Fatty Acids | 0.87 (0.78, 0.97) | 0.012 | 0.64 (0.51, 0.81) | <0.001 |
| 23450 | Docosahexaenoic Acid | 0.19 (0.06, 0.59) | 0.004 | 0.01 (0.00, 0.09) | <0.001 |
| 23478 | Creatinine | 40.01 (1.13, 1412.99) | 0.043 | 28.75 (0.01, 72111.74) | 0.400 |
| 23479 | Albumin | 0.94 (0.91, 0.96) | <0.001 | 0.91 (0.86, 0.95) | <0.001 |
| 23480 | Glycoprotein Acetyls | 4.53 (2.17, 9.45) | <0.001 | 3.83 (0.88, 16.55) | 0.073 |
| 23523 | Concentration of IDL Particles | 0.00 (0.00, 0.00) | 0.013 | 0.00 (0.00, 0.00) | <0.001 |
| 23524 | Total Lipids in IDL | 0.60 (0.43, 0.82) | 0.001 | 0.19 (0.10, 0.37) | <0.001 |
| 23525 | Phospholipids in IDL | 0.17 (0.04, 0.67) | 0.011 | 0.00 (0.00, 0.03) | <0.001 |
| 23528 | Free Cholesterol in IDL | 0.08 (0.01, 0.38) | 0.002 | 0.00 (0.00, 0.01) | <0.001 |
| 23534 | Cholesteryl Esters in Large LDL | 0.55 (0.36, 0.84) | 0.005 | 0.14 (0.06, 0.33) | <0.001 |
| 23535 | Free Cholesterol in Large LDL | 0.14 (0.04, 0.44) | 0.001 | 0.00 (0.00, 0.03) | <0.001 |
| 23538 | Total Lipids in Medium LDL | 0.55 (0.33, 0.94) | 0.028 | 0.11 (0.04, 0.34) | <0.001 |
| 23540 | Cholesterol in Medium LDL | 0.42 (0.20, 0.88) | 0.021 | 0.04 (0.01, 0.21) | <0.001 |
| 23541 | Cholesteryl Esters in Medium LDL | 0.34 (0.13, 0.94) | 0.036 | 0.02 (0.00, 0.15) | <0.001 |
| 23546 | Phospholipids in Small LDL | 0.01 (0.00, 0.53) | 0.026 | 0.00 (0.00, 0.00) | 0.001 |
| 23547 | Cholesterol in Small LDL | 0.10 (0.01, 0.73) | 0.023 | 0.00 (0.00, 0.02) | <0.001 |
| 23548 | Cholesteryl Esters in Small LDL | 0.06 (0.00, 0.86) | 0.038 | 0.00 (0.00, 0.01) | <0.001 |
| 23549 | Free Cholesterol in Small LDL | 0.00 (0.00, 0.12) | 0.010 | 0.00 (0.00, 0.00) | <0.001 |
| 23551 | Concentration of Very Large HDL Particles | 0.00 (0.00, 0.00) | 0.001 | 0.00 (0.00, 0.00) | 0.012 |
| 23555 | Cholesteryl Esters in Very Large HDL | 0.00 (0.00, 0.04) | <0.001 | 0.00 (0.00, 0.31) | 0.025 |
| 23563 | Free Cholesterol in Large HDL | 0.00 (0.00, 0.08) | <0.001 | 0.00 (0.00, 0.19) | 0.013 |
| 23565 | Concentration of Medium HDL Particles | 0.00 (0.00, 0.00) | <0.001 | 0.00 (0.00, 0.00) | 0.001 |
| 23566 | Total Lipids in Medium HDL | 0.48 (0.31, 0.74) | 0.001 | 0.26 (0.11, 0.63) | 0.003 |
| 23567 | Phospholipids in Medium HDL | 0.23 (0.09, 0.59) | 0.002 | 0.06 (0.01, 0.43) | 0.005 |
| 23575 | Cholesterol in Small HDL | 0.14 (0.03, 0.58) | 0.007 | 0.01 (0.00, 0.15) | 0.001 |
| 23577 | Free Cholesterol in Small HDL | 0.00 (0.00, 0.06) | 0.003 | 0.00 (0.00, 0.00) | <0.001 |

^a^ Adjusted for age, sex, Townsend deprivation index, physical activity, smoking status, BMI, total energy, and total sugar intake. HR refers to the hazard ratio of IBD or CD as per one-standard deviation increase in metabolite levels.

CD, Crohn’s disease; CI, confidence interval; DHA, docosahexaenoic acid; IBD, inflammatory bowel disease; HDL, high-density lipoprotein; HR, hazard ratio; IDL, intermediate density lipoprotein; LDL, low-density lipoprotein; PUFA, polyunsaturated fatty acids; UKB, UK Biobank; WGCNA, weighted gene co-expression network analysis.

### **Table S15.** Sensitivity analysis of associations between UPF metabolic signature and incident CD, additionally adjusted for C-reactive protein level and INFLA-score

| **Exposure** | IBD |  |  | CD |  |  |
| --- | --- | --- | --- | --- | --- | --- |
|  | Cases | HR (95% CI) ^a^ | *P* | Cases | HR (95% CI) | *P* |
| **Additionally adjusted for C-reactive protein level** | | | | | | |
| **Discovery cohort** | | | | | | |
| Per SD |  | **1.07 (1.03, 1.11)** | **0.001** |  | **1.11 (1.02, 1.22)** | **0.020** |
| Low | 13 | Ref |  | 1 | Ref |  |
| High | 39 | **2.57 (1.29, 5.14)** | **0.007** | 11 | 4.74 (0.57, 39.30) | 0.149 |
| **Internal validation** | | | | | | |
| Per SD |  | **1.20 (1.10, 1.31)** | **<0.001** |  | **1.34 (1.16, 1.56)** | **<0.001** |
| T1 (lowest) | 124 | Ref |  | 28 | Ref |  |
| T2 | 188 | **1.48 (1.17, 1.88)** | **0.001** | 44 | 1.63 (1.00, 2.65) | 0.050 |
| T3 (highest) | 215 | **1.62 (1.24, 2.11)** | **<0.001** | 63 | **2.42 (1.43, 4.10)** | **0.001** |
| *P* for trend |  |  | **<0.001** |  |  | **0.001** |
| **Additionally adjusted for INFLA-score** | | | | | | |
| **Discovery cohort** | | | | | | |
| Per SD |  | **1.07 (1.03, 1.11)** | **0.001** |  | **1.11 (1.02, 1.22)** | **0.018** |
| Low | 13 | Ref |  | 1 | Ref |  |
| High | 39 | **2.53 (1.26, 5.06)** | **0.009** | 11 | 4.93 (0.59, 40.81) | 0.139 |
| **Internal validation** | | | | | | |
| Per SD |  | **1.18 (1.08, 1.30)** | **<0.001** |  | **1.34 (1.15, 1.55)** | **<0.001** |
| T1 (lowest) | 124 | Ref |  | 28 | Ref |  |
| T2 | 188 | **1.50 (1.17, 1.91)** | **0.001** | 44 | **1.67 (1.01, 2.76)** | **0.045** |
| T3 (highest) | 215 | **1.58 (1.20, 2.07)** | **0.001** | 63 | **2.43 (1.40, 4.20)** | **0.002** |
| *P* for trend |  |  | **0.002** |  |  | **0.001** |

^a^ The fully-adjusted model was adjusted for age, sex, Townsend deprivation index, physical activity, smoking status, BMI, total energy, and total sugar intake.

CD, Crohn’s disease; CI, confidence interval; HR, hazard ratio; IBD, inflammatory bowel disease; SD, standard deviation; T1 to T3, tertile 1 to tertile 3; UPF, ultra-processed food.

### **Table S16.** Sensitivity analysis of associations between UPF metabolic signature and incident CD, excluding incident cases in the first 2 years of follow-up ^a^

| **Exposure** | IBD |  |  | CD |  |  |
| --- | --- | --- | --- | --- | --- | --- |
|  | Cases | HR (95% CI) | *P* | Cases | HR (95% CI) | *P* |
| **Discovery cohort (n=10,193)** | | | | | | |
| Per SD |  | **1.06 (1.01, 1.11)** | **0.01** |  | 1.08 (0.98, 1.19) | 0.104 |
| Low | 13 | Ref |  | 1 | Ref |  |
| High | 39 | **2.19 (1.04, 4.61)** | **0.038** | 11 | 3.67 (0.43, 31.52) | 0.237 |
| **Internal validation (n=90,526)** | | | | | | |
| Per SD |  | **1.22 (1.11, 1.34)** | **<0.001** |  | **1.36 (1.17, 1.58)** | **<0.001** |
| T1 (lowest) | 94 | Ref |  | 23 | Ref |  |
| T2 | 155 | **1.54 (1.19, 1.99)** | **0.001** | 37 | 1.67 (0.98, 2.86) | 0.061 |
| T3 (highest) | 166 | **1.73 (1.29, 2.30)** | **<0.001** | 53 | **2.54 (1.43, 4.52)** | **0.002** |
| *P* for trend |  |  | **<0.001** |  |  | **0.001** |

^a^ Adjusted for age, sex, Townsend deprivation index, physical activity, smoking status, BMI, total energy, and total sugar intake.

CD, Crohn’s disease; CI, confidence interval; HR, hazard ratio; IBD, inflammatory bowel disease; SD, standard deviation; T1 to T3, tertile 1 to tertile 3; UPF, ultra-processed food.

### **Table S17.** Correlation coefficients between the levels of circulating metabolites, ultra-processed food intake, and CD-related outcomes in the ONE-IBD cohort

|  | Spearman correlation | | | |
| --- | --- | --- | --- | --- |
|  | Spearman ρ | *P* | Spearman ρ | *P* |
|  | Association with UPF intake in healthy controls | | Association with UPF intake in IBD participants | |
| DHA | **-0.16** | **0.003** | **-0.25** | **<0.001** |
| Creatinine | <0.01 | 0.946 | -0.08 | 0.170 |
| Lactate | 0.06 | 0.512 | -0.11 | 0.053 |
| Acetate | -0.01 | 0.863 | 0.10 | 0.075 |
| Glucose | -0.05 | 0.351 | 0.04 | 0.482 |
| Pyruvate | **0.12** | **0.027** | **-0.15** | **0.009** |
| Linoleic acid | -0.01 | 0.986 | **-0.13** | **0.026** |
|  | Association with UPF intake in CD participants | | Association with CRP in CD participants | |
| DHA | **-0.28** | **<0.001** | **-0.26** | **0.032** |
| Creatinine | 0.05 | 0.538 | -0.22 | 0.071 |
| Lactate | 0.01 | 0.933 | -0.13 | 0.286 |
| Acetate | 0.01 | 0.975 | 0.10 | 0.419 |
| Glucose | 0.01 | 0.902 | 0.15 | 0.228 |
| Pyruvate | -0.09 | 0.234 | -0.06 | 0.645 |
| Linoleic acid | -0.08 | 0.328 | -0.11 | 0.365 |
|  | Association with CDAI in CD participants | | Association with biological relapse in CD participants | |
| DHA | **-0.24** | **<0.001** | **-0.19** | **0.015** |
| Creatinine | -0.04 | 0.472 | -0.13 | 0.105 |
| Lactate | -0.06 | 0.269 | 0.03 | 0.725 |
| Acetate | 0.05 | 0.301 | 0.00 | 0.972 |
| Glucose | 0.05 | 0.301 | -0.05 | 0.521 |
| Pyruvate | **-0.15** | **0.004** | -0.11 | 0.178 |
| Linoleic acid | **-0.15** | **0.003** | -0.12 | 0.149 |

CD, Crohn’s disease; CDAI, Crohn’s disease activity index; CI, confidence interval; DHA, docosahexaenoic acid; IBD, inflammatory bowel disease; OR, odds ratio; SD, standard deviation; UPF, ultra-processed food. Biological relapse refers to a C-reactive protein level over 10 mg/L during the follow-up.

### **Table S18.** Odds ratios of the levels of circulating metabolites significantly associated with risks of CD in the ONE-IBD cohort

|  | Crude model |  | Adjusted model ^a^ |  |
| --- | --- | --- | --- | --- |
|  | OR (95% CI) | *P* | OR (95% CI) | *P* |
| DHA |  |  |  |  |
| Per SD | **0.21 (0.16, 0.28)** | **<0.001** | **0.23 (0.17, 0.30)** | **<0.001** |
| High vs low | **0.12 (0.09, 0.18)** | **<0.001** | **0.15 (0.10, 0.21)** | **<0.001** |
| Tertile 1 | Ref |  | Ref |  |
| Tertile 2 | **0.31 (0.21, 0.47)** | **<0.001** | **0.32 (0.21, 0.49)** | **<0.001** |
| Tertile 3 | **0.05 (0.03, 0.08)** | **<0.001** | **0.05 (0.03, 0.09)** | **<0.001** |
| Lactate |  |  |  |  |
| Per SD | **1.00 (1.00, 1.00)** | **<0.001** | **1.00 (1.00, 1.00)** | **<0.001** |
| High vs low | **12.95 (9.00, 18.89)** | **<0.001** | **10.01 (6.86, 14.79)** | **<0.001** |
| Tertile 1 | Ref |  | Ref |  |
| Tertile 2 | **4.04 (2.56, 6.50)** | **<0.001** | **3.11 (1.93, 5.08)** | **<0.001** |
| Tertile 3 | **35.66 (21.33, 61.64)** | **<0.001** | **24.71 (14.54, 43.28)** | **<0.001** |
| Glucose |  |  |  |  |
| Per SD | **1.00 (1.00, 1.00)** | **<0.001** | **1.00 (1.00, 1.00)** | **<0.001** |
| High vs low | **0.10 (0.07, 0.14)** | **<0.001** | **0.10 (0.06, 0.14)** | **<0.001** |
| Tertile 1 | Ref |  | Ref |  |
| Tertile 2 | **0.09 (0.05, 0.14)** | **<0.001** | **0.07 (0.06, 0.16)** | **<0.001** |
| Tertile 3 | **0.03 (0.02, 0.05)** | **<0.001** | **0.04 (0.02, 0.06)** | **<0.001** |
| Pyruvate |  |  |  |  |
| Per SD | **1.02 (1.01, 1.02)** | **<0.001** | **1.02 (1.01, 1.02)** | **<0.001** |
| High vs low | **3.53 (2.57, 4.87)** | **0.001** | **3.55 (2.52, 5.04)** | **<0.001** |
| Tertile 1 | Ref |  | Ref |  |
| Tertile 2 | **4.29 (2.86, 6.52)** | **<0.001** | **4.43 (2.86, 6.97)** | **<0.001** |
| Tertile 3 | **5.77 (3.82, 8.81)** | **<0.001** | **6.07 (3.90, 9.61)** | **<0.001** |
| Linoleic acid | |  |  |  |
| Per SD | **0.99 (0.99, 1.00)** | **0.001** | **0.99 (0.99, 1.00)** | **0.002** |
| High vs low | **0.56 (0.41, 0.75)** | **<0.001** | **0.60 (0.43, 0.83)** | **0.002** |
| Tertile 1 | Ref |  | Ref |  |
| Tertile 2 | **0.52 (0.36, 0.76)** | **0.012** | **0.55 (0.37, 0.83)** | **0.004** |
| Tertile 3 | **0.42 (0.28, 0.61)** | **<0.001** | **0.45 (0.3, 0.67)** | **<0.001** |

^a^ The adjusted model was adjusted for age and sex.

CD, Crohn’s disease; CI, confidence interval; DHA, docosahexaenoic acid; OR, odds ratio; SD, standard deviation; UPF, ultra-processed food.

### **Table S19.** Mendelian randomization and colocalization analysis for associations of DHA with IBD and CD

| Outcome | Exposure | No. of IV | Inverse variant weighing | | | Colocalization | | | Hyprcolocalization | | | |
| --- | --- | --- | --- | --- | --- | --- | --- | --- | --- | --- | --- | --- |
|  |  |  | OR (95% CI) | *P* | *P* for Q-test | Lead SNP | Gene | PPH4 | Traits | Colocalized SNP (consequence) | Gene | PPFC (PPE) |
| IBD | DHA | 39 | 0.72 (0.63, 0.82) | 1.37E-10 | 2.97E-03 | rs174546 | *FADS1* (chr 11) | 96.30% | DHA, IBD, CD | rs174546 (3 Prime UTR Variant) | *FADS1* (chr 11) | 0.7365 (0.3479) |
| CD | DHA | 39 | 0.70 (0.60, 0.81) | 1.38E-05 | 1.07E-02 |  |  | 95.80% |  |  |  |  |

CD, Crohn’s disease; CI, confidence interval; DHA, docosahexaenoic acid; IBD, inflammatory bowel disease; IV, instrumental variable; OR, odds ratio; SNP, single nucleotide polymorphism; PPFC, posterior probability of full colocalization; PPE, proportion of PPFC explained by the listed SNP; PPH4, posterior probability of hypothesis 4; UTR, untranslated region.

### **Table S20.** Mendelian randomization sensitivity analysis for associations of DHA with IBD and CD

| Outcome | Exposure | No. of IV | Weighted median | | MR Egger | | | MR PRESSO | | |
| --- | --- | --- | --- | --- | --- | --- | --- | --- | --- | --- |
|  |  |  | OR (95% CI) | *P* | OR (95% CI) | *P* | *P* for pleiotropy | OR (95% CI) | *P* | Number of outliers |
| IBD | DHA | 39 | 0.78 (0.71, 0.85) | 1.28E-08 | 0.77 (0.69, 0.86) | 4.25E-05 | 7.93E-01 | 0.78 (0.72, 0.84) | 1.52E-07 | 0 |
| CD | DHA | 39 | 0.69 (0.60, 0.80) | 7.96E-07 | 0.72 (0.58, 0.90) | 5.94E-03 | 8.82E-01 | 0.73 (0.63, 0.84) | 6.75E-05 | 1 |

CD, Crohn’s disease; CI, confidence interval; DHA, docosahexaenoic acid; IBD, inflammatory bowel disease; IV, instrumental variable; OR, odds ratio.

### **Table S21.** Pair-Wise Conditional analysis and Colocalization analysis for DHA and CD

| Exposure | Outcome | SNP in exposure data ^a^ | SNP in outcome data | Number of SNPs | H0 | H1 | H2 | H3 | H4 | Log_abf_all ^b^ |
| --- | --- | --- | --- | --- | --- | --- | --- | --- | --- | --- |
| DHA QTL | CD | unconditioned | unconditioned | 4417 | 0.00E+00 | 0.000856 | 0.00E+00 | 0.037072 | 0.962072 | 1540.62 |
| DHA QTL | CD | rs174546 | unconditioned | 1149 | 2.97E-42 | 0.002047 | 1.28E-40 | 0.087252 | 0.910701 | 95.6192 |
| DHA QTL | CD | rs174565 | unconditioned | 1119 | 7.50E-09 | 0.124516 | 3.58E-08 | 0.594554 | 0.28093 | 18.7084 |
| DHA QTL | CD | rs2524299 | unconditioned | 1128 | 2.02E-42 | 0.137316 | 1.02E-41 | 0.691131 | 0.171553 | 96.007 |
| DHA QTL | CD | rs174602 | unconditioned | 1108 | 1.03E-07 | 0.159512 | 4.86E-07 | 0.752258 | 0.088229 | 16.0883 |
| DHA QTL | CD | rs2238001 | unconditioned | 1104 | 3.03E-06 | 0.160211 | 1.43E-05 | 0.75535 | 0.084422 | 12.7063 |
| DHA QTL | CD | rs174603 | unconditioned | 1109 | 3.26E-17 | 0.166397 | 1.54E-16 | 0.784748 | 0.048855 | 37.9629 |
| DHA QTL | CD | rs198448 | unconditioned | 1107 | 6.89E-07 | 0.170206 | 3.25E-06 | 0.802577 | 0.027213 | 14.1887 |
| DHA QTL | CD | rs7394579 | unconditioned | 1117 | 1.46E-08 | 0.171102 | 6.89E-08 | 0.808575 | 0.020323 | 18.0437 |
| DHA QTL | CD | rs11539526 | unconditioned | 1127 | 1.28E-17 | 0.171351 | 6.04E-17 | 0.808572 | 0.020077 | 38.8976 |
| DHA QTL | CD | rs6591657 | unconditioned | 1112 | 2.16E-07 | 0.172615 | 1.02E-06 | 0.814027 | 0.013357 | 15.3476 |
| DHA QTL | CD | rs174620 | unconditioned | 1119 | 2.04E-05 | 0.173089 | 9.61E-05 | 0.816334 | 0.01046 | 10.8016 |
| DHA QTL | CD | rs3781973 | unconditioned | 1108 | 5.31E-09 | 0.173399 | 2.50E-08 | 0.817557 | 0.009044 | 19.0538 |
| DHA QTL | CD | rs11230735 | unconditioned | 1104 | 3.85E-05 | 0.173418 | 1.82E-04 | 0.817708 | 0.008653 | 10.1644 |
| DHA QTL | CD | rs12796595 | unconditioned | 1108 | 2.73E-08 | 0.173975 | 1.29E-07 | 0.820349 | 0.005676 | 17.4161 |
| DHA QTL | CD | rs441938 | unconditioned | 1104 | 1.04E-07 | 0.17405 | 4.91E-07 | 0.82069 | 0.005259 | 16.0776 |
| DHA QTL | CD | rs174473 | unconditioned | 1104 | 4.30E-24 | 0.174187 | 2.03E-23 | 0.821278 | 0.004535 | 53.804 |
| DHA QTL | CD | rs3741259 | unconditioned | 1104 | 3.52E-07 | 0.174331 | 1.66E-06 | 0.822015 | 0.003653 | 14.8587 |
| DHA QTL | CD | rs4963279 | unconditioned | 1104 | 4.43E-15 | 0.17453 | 2.09E-14 | 0.822954 | 0.002516 | 33.05 |
| DHA QTL | CD | rs259872 | unconditioned | 1104 | 1.61E-17 | 0.174535 | 7.59E-17 | 0.822978 | 0.002487 | 38.6685 |
| DHA QTL | CD | rs259877 | unconditioned | 1105 | 6.61E-22 | 0.174539 | 3.12E-21 | 0.822999 | 0.002462 | 48.7679 |
| DHA QTL | CD | rs169167 | unconditioned | 1110 | 7.37E-05 | 0.174493 | 3.47E-04 | 0.822724 | 0.002362 | 9.51586 |
| DHA QTL | CD | rs3018732 | unconditioned | 1109 | 6.73E-12 | 0.174577 | 3.17E-11 | 0.823092 | 0.002331 | 25.7246 |

^a^ Note: when a column contains a SNP, this should be read that that SNP was not conditioned upon to generate that colocalization result. So, when reading the row with rs174546, this shows that the remaining SNPs in the table were all conditioned upon.

^b^ Log_abf_all represents the log approximate Bayes factor quantifying the joint statistical evidence for colocalization across all SNPs under a given conditional configuration, as estimated within the SuSiE-based colocalization framework.

CD, Crohn’s disease; DHA, docosahexaenoic acid; SNP, single nucleotide polymorphism.
